# Evaluation of Precuneus and Fusiform Gyrus-Based Radiomic biomarkers for Alzheimer’s disease Classification and Progression

**DOI:** 10.1101/2024.05.15.24307407

**Authors:** Ishsirjan Kaur Chandok, Simran Sharma, Kavita Kundal, Sushree Sangita Kar, Rehan Khan, Arunabha Majumdar, Neeraj Kumar, Rahul Kumar

**Affiliations:** Department of Biotechnology, Indian Institute of Technology Hyderabad, Kandi, Telangana 502285, India; Department of Mathematics, Indian Institute of Technology Hyderabad, Kandi, Telangana 502285, India; Department of Liberal Arts, Indian Institute of Technology Hyderabad, Kandi, Telangana 502285, India

**Keywords:** MRI, Alzheimer’s Disease, Fusiform, Precuneus, Early imaging biomarkers

## Abstract

Alzheimer’s disease (AD) is characterized by progressive neurodegeneration, with early structural changes detectable in specific brain regions. This study explores the diagnostic and prognostic utility of MRI-derived radiomic features from the precuneus and fusiform gyrus for identifying and tracking AD progression. T1-weighted MRI scans from 382 participants; 134 cognitively normal (CN), 149 with mild cognitive impairment (MCI), and 99 with AD were analyzed across four time points (0, 6, 12, and 24 months). Using the FreeSurfer automated pipeline, nine radiomic features were extracted bilaterally from the precuneus and fusiform gyrus. Statistical comparisons were conducted using the Mann-Whitney U test with Benjamini–Hochberg correction. Diagnostic classification was performed using random forest models, while disease progression was modeled using multiple linear regression and ARIMA-based time-series approaches.Significant reductions in gray matter volume (GMV) and cortical thickness (CT) were observed in AD patients compared to CN and MCI groups (Padj < 0.001). Random forest classifiers achieved high training accuracies: 98.21% (AD vs. CN), 96.98% (AD vs. MCI), and 99.31% (MCI vs. CN). Prognostic modeling showed highest predictive performance in the left fusiform gyrus (GMV: r = 0.97, CT: r = 0.93), followed by the left precuneus, right fusiform, and right precuneus. Time-series models outperformed linear regression in most cases, reinforcing temporal consistency in radiomic progression.Radiomic features from the precuneus and fusiform gyrus enable robust classification of AD stages and accurate modeling of disease progression. These regions represent promising non-invasive biomarkers for early diagnosis and longitudinal monitoring of Alzheimer’s disease.

## Introduction

Alzheimer’s disease (AD) is the primary cause of dementia, affecting 60% to 80% of cases globally. It’s characterized by memory loss, cognitive decline, and behavioral changes, leading to a loss of independence. The pathological progression of AD is typically described in three stages: a preclinical phase in which individuals are cognitively normal, followed by mild cognitive impairment (MCI), and culminating in overt dementia.^1^Mild cognitive impairment (MCI) is a transitional stage, with about 35% of cases progressing to AD or dementia within 3-4 years.^1^ In 2015, AD cost around 818 billion USD globally, impacting over 47 million individuals^2^. By 2050, its prevalence is projected to quadruple, affecting nearly 1 in 85 people, primarily due to population aging^3^.

In AD, the precuneus and fusiform gyrus are pivotal regions implicated in the neurodegenerative cascade leading to cognitive impairment and functional decline. The precuneus serves as a central hub for various cognitive processes, including episodic memory retrieval, visuospatial processing, and self-referential cognition.^4–8^ Numerous neuroimaging studies have consistently revealed significant atrophy, hypometabolism within the precuneus, and reduced connectivity with the default mode network in individuals with AD^9–14^, particularly in the early stages of the disease ^15^. The atrophy in the precuneus area is also shown even in the non-amnestic Aβneg-AD individuals ^16^.

Conversely, the fusiform gyrus is renowned for its specialization in facial recognition and the processing of complex visual stimuli ^17^. In AD, alterations in the fusiform gyrus have been observed as reduced GMV^18^ and disrupted functional connectivity^19^. These changes underlie the characteristic deficits in facial recognition and discrimination encountered by AD patients, contributing to their impaired social cognition and interpersonal interactions.

Emerging evidence suggests that the precuneus serves as a convergence zone integrating sensory, mnemonic, and attentional information critical for cognitive processing, including facial recognition mediated by the fusiform gyrus ^8,20–23^. Disruptions within this network, specifically in face perception, may lead to the cascade of cognitive impairments like disruption in face-related episodic memory retrieval, limited interpersonal relations, and poor spatial awareness observed in AD, encompassing deficits in memory consolidation, attentional control, and social cognition. Understanding of the structural and functional alterations within these regions may offer valuable biomarkers for early diagnosis and monitoring disease progression in AD. The present study employs a machine learning-based approach leveraging radiomics analysis of MRI data to identify novel biomarkers from the precuneus and fusiform gyrus by systematically analyzing volumetric and surface features extracted from MRI images of individuals across different stages of cognitive impairment, including cognitively normal (CN), mild cognitive impairment (MCI), and AD. We have developed an ML-based classifier that can categorize individuals into MCI or AD based on their precuneus and fusiform gyrus features.

Furthermore, we utilized these radiomic features in a linear regression and time-series model to quantitatively analyze and predict the progression of AD, providing valuable insights into the disease trajectory. Figure 1 depicts the overall schema of this study

**Figure 1:**
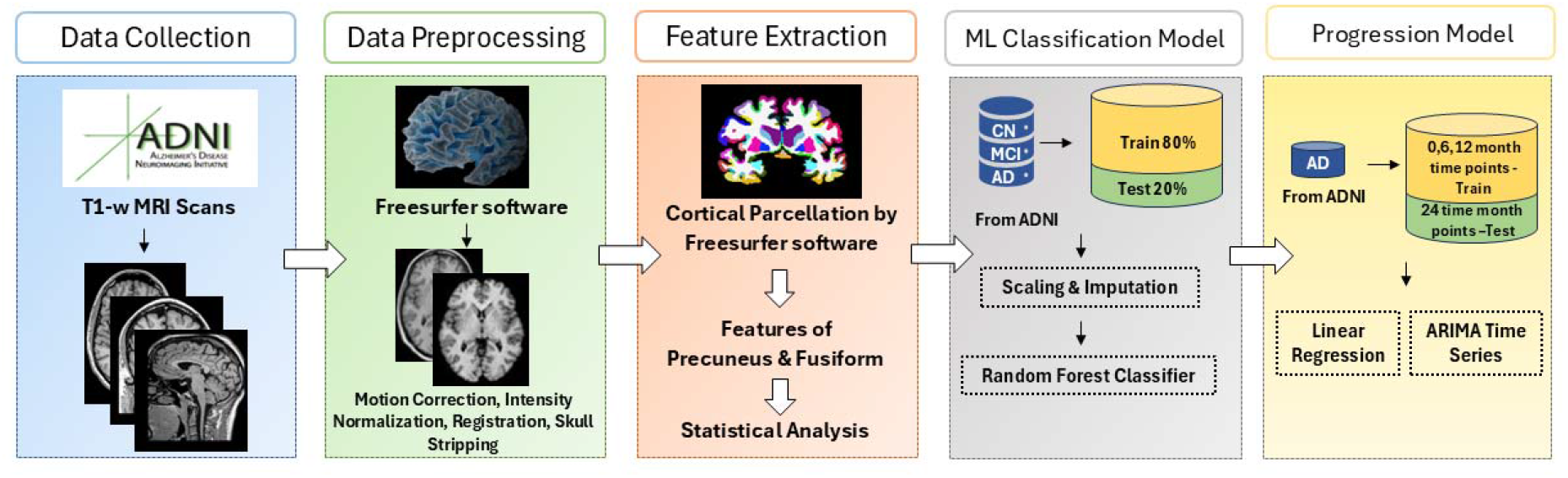
Schematic diagram showing the study plan.

## Methodology

### Data Description

We downloaded the T1-weighted MRI images from the ADNI1 collection (named ADNI1: Complete 3yr 1.5T) in the Alzheimer’s Disease Neuroimaging Initiative (ADNI) database, which comprises cohorts with 134 CN, 149 MCI, and 99 AD subjects. The age range for CN individuals spans from 60 to 92 years while those with MCI range from 55 to 90 years, and for those diagnosed with AD, it ranges from 56 to 91 years. In the ADNI dataset, Mini-Mental State Examination (MMSE), Clinical Dementia Rating (CDR), and the National Institute of Neurological and Communicative Disorders and Stroke/Alzheimer’s Disease and Related Disorders Association (NINCDS/ADRDA, ^24^ were the criteria used to assess the level of cognition in the samples (CN, MCI, and AD). CNs have an MMSE score in the range of 24-30, a CDR score of 0, and no signs of depression. MCIs have an MMSE score in the range of 24-30 and a CDR score of 0.5, with memory complaints and minimally impaired daily activities. AD criteria include MMSE scores of 20-26 (inclusive) and a CDR of 0.5 or 1.0, fulfilling NINCDS/ADRDA criteria. For each subject, four different time points were available i.e., 0-month, 6-month, 12-month and 24-month. In the ADNI collection the 1.5T MRI images were acquired with multicoil phased-array head coil (MA), FOV set at 240 × 240 mm, flip angle of 8 degrees, and TR of 2300ms ^25^.Images were pre-processed for multiplanar reconstruction-reslicing (MPR-R), B1 radiofrequency pulse for non-uniformity correction, gradient warping, and N3 to reduce intensity non-uniformity^26^

### Radiomic-Based Feature Extraction and Statistical Method

We implemented the fully automated ‘Recon-All’ pipeline provided by FreeSurfer software for feature extraction ^27^. It is an open-source package to analyze and display structural, functional, and diffusion neuroimaging data from cross-sectional and longitudinal research This pipeline initially follows a few image processing steps, i.e., motion artifact correction, transformation to Talairach image space, intensity normalization, and skull stripping ^28^. Cortical parcellation was performed for feature extraction using the Desikan-Killiany atlas, which divides the MRI scans into 34 discrete anatomical regions^29^. 9 different types of radiomic features i.e., the total gray matter volume (GMV, mm^3^), the number of vertices, the average cortical thickness (CT, mm), the standard cortical thickness (mm), the total surface areas (mm^2^), the integrated rectified mean curvature (mm^-1^), the integrated rectified Gaussian curvature (mm^-2^), the folding index, and the intrinsic curvature index) were calculated for the precuneus and fusiform gyrus from both left and right hemispheres. We applied the nonparametric Mann-Whitney U test to compare these nine features of the precuneus and fusiform gyrus between CN, MCI, and AD patients at four time points. The p-values were adjusted using the Benjamin-Hochberg correction method. Box and whisker plots were used to provide a graphical representation of the distribution of these groups.

### Longitudinal Machine Learning Predictive Model Development

#### 1. Classification model

We combined above mentioned 9 radiomic features of both right and left hemispheres of precuneus and fusiform gyrus (total 36 features) and developed three binary classification models, to classify AD vs CN, MCI vs AD, and CN vs MCI. We also used age of the individuals as one of the features to evaluate its impact on the performance of these models. Additionally, classification models that solely used precuneus or fusiform based features were also constructed. We employed the random Forest Classifier (RFC) algorithm to develop the classification models. First, the dataset was divided into a training dataset (80%) and a test dataset (20%), and the KNN imputation method was implemented to ensure uniformity^30^. The class imbalance was addressed through random under sampling, while hyperparameter optimization of the RFC was carried out using the “RandomizedSearchCV” function to enhance model performance. The model performance was evaluated using various statistical metrics, i.e., accuracy,precision, recall, F1-score, ROC AUC, and Matthews Correlation Coefficient (MCC) ^31– 33^.

#### 2. Linear regression and Time-series model

##### Model 1: Multiple Linear Regression

Multiple linear regression is used to predict the temporal evolution of each brain feature. Each feature is examined separately, and its value at 24 months is estimated as a linear function of its values at the three prior time points (*t =* 0, 1, 2).

###### Model Formulation

We assume the following linear model for each feature *j*:

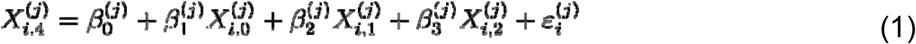

Where:

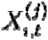is the value of the feature *j* for the individual *i* at the time *t*.

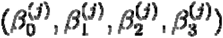 re the *j-*th feature-specific regression coefficients. These are the intercept coefficient and slope coefficients corresponding to the three consecutive time points, respectively.

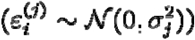 denotes the random error term, assumed i.i.d. Gaussian..

Ordinary least squares (OLS) are used to estimate this model by minimizing the residual sum of squares:

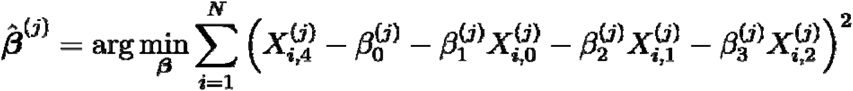

Using the OLS estimators, for a given feature *j*, the predicted value is:

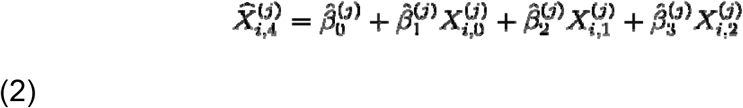

##### Model 2: Time-Series Modelling with ARIMA

For the analysis and prediction of time-series data, the Autoregressive Integrated Moving Average (ARIMA) model is an effective tool. Three components are used to form an ARIMA(*p,d,q*) model:

###### Autoregressive (AR) of order *p*

This component models the current value as a linear combination of its previous values:

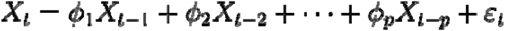

Where *ε*_*t*_ is a white noise error term.

###### Integrated (I) of order *d*

This component accounts for non-stationarity by differencing the series *d* times:

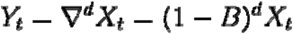

###### Moving Average (MA) of order *q*

This component models the current value as a linear combination of past errors:

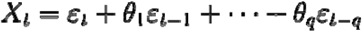

### ARIMA Model Definition

An ARIMA(*p,d,q*) model is fitted to each subject’s time-series. The general form is:

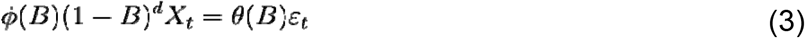

Where *B* is the backshift operator, and □(*B*), θ(*B*) are polynomials that represent the moving average and autoregressive components, respectively.

For the subject *i*, let *X*_*i,l*_ represent the measurement at time *t*. Based on the known values at *t=*0, 1, 2, 4, cubic spline interpolation is used to estimate the 18-month data (*t*=3) as it is not observed. We then form a complete time-series for each subject:

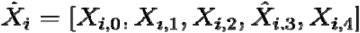

The series 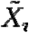 for each subject is fitted to an ARIMA(*p, d, q*) model. The auto.arima() function in R is used to automatically choose the model order (*p, d, q*) using AIC.

### Model Evaluation Metrics

For both modelling approaches (regression and ARIMA), the following performance metrics are used to evaluate predictions at *t* = 24 months:

### Pearson Correlation Coefficient (r)

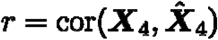

### Root Mean Squared Error (RMSE)

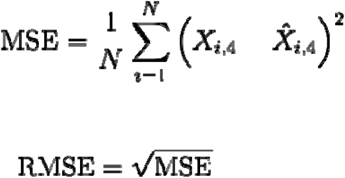

### Mean Absolute Percentage Error (MAPE)

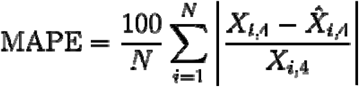

These measurements offer a thorough comparison of the accuracy of the two modelling frameworks in predicting the evaluation of brain features. Longitudinal brain feature trajectories are modelled using both MLR and ARIMA. Although ARIMA delivers individualized time-series modelling that incorporates subject-specific temporal dynamics and autocorrelation, MLR provides an overall linear relationship across subjects. The applicability of each strategy for various kinds of radiomic patterns can be evaluated by comparing their predictive performance.

## Results

### Statistical Analysis for MRI-based Radiomic Features

We extracted the volumetric and surface related features from MRI images and applied Mann-Whitney U test to compare these nine radiomic features of precuneus and fusiform gyrus for right and left hemisphere (Figure 2-3, Supplementary Table 1 and Supplementary Figures 1-16). Among these features, GMV showed significant reduction in fusiform gyrus among AD patients compared to MCI and CN at all 4 timepoint 0,6,12 and 24-months (Padj<0.05) Interestingly, the GMV of the precuneus was also significantly reduced in AD patients as compared to MCI and CN at all time points (*Padj<0*.*05*)(Figure 2 and Supplementary Table 1). In addition to GMV, we also observed significant reduction in CT in both precuneus and fusiform gyrus (Supplementary Table 1 and Figure 3). We also performed statistical analysis for other radiomic features and observed significant differences (Supplementary Table 1 and Supplementary Figures 1-16). Additionally, at the 24-month time point, boxplots illustrated continued decline in GMV and CT in the right fusiform gyrus and precuneus among AD patients (Figure 4 and Supplementary Figure 17).

**Figure 2:**
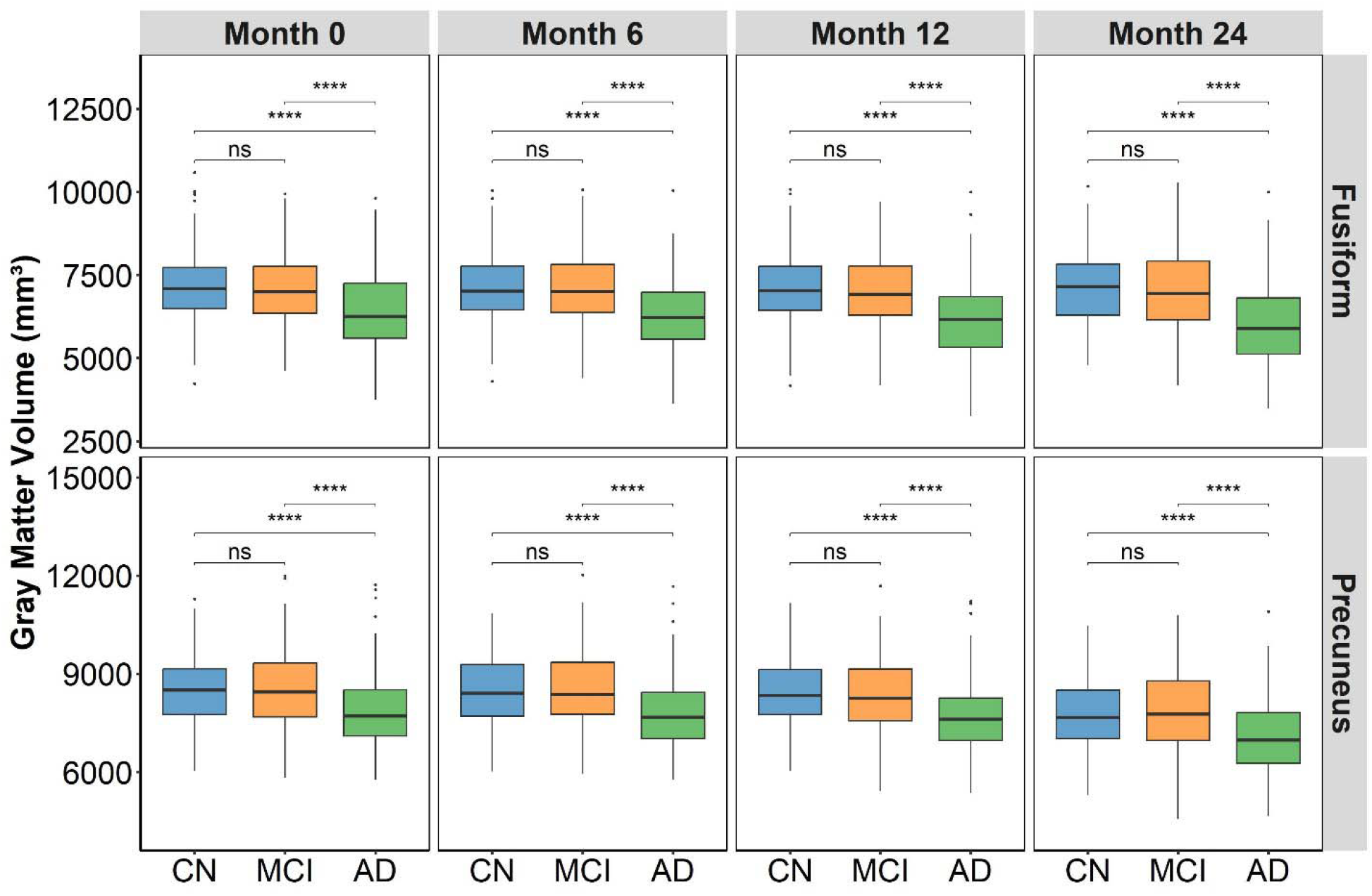
Boxplots illustrating GMV of the fusiform gyrus and precuneus (right hemisphere) in CN, MCI, and AD individuals at 0-month, 6-month, 12-month and 24-month time point, indicating significant reductions in AD. Statistical comparisons are indicated (*: p < 0.05; **: p < 0.01; ***: p < 0.001; ****: p < 0.0001).

**Figure 3:**
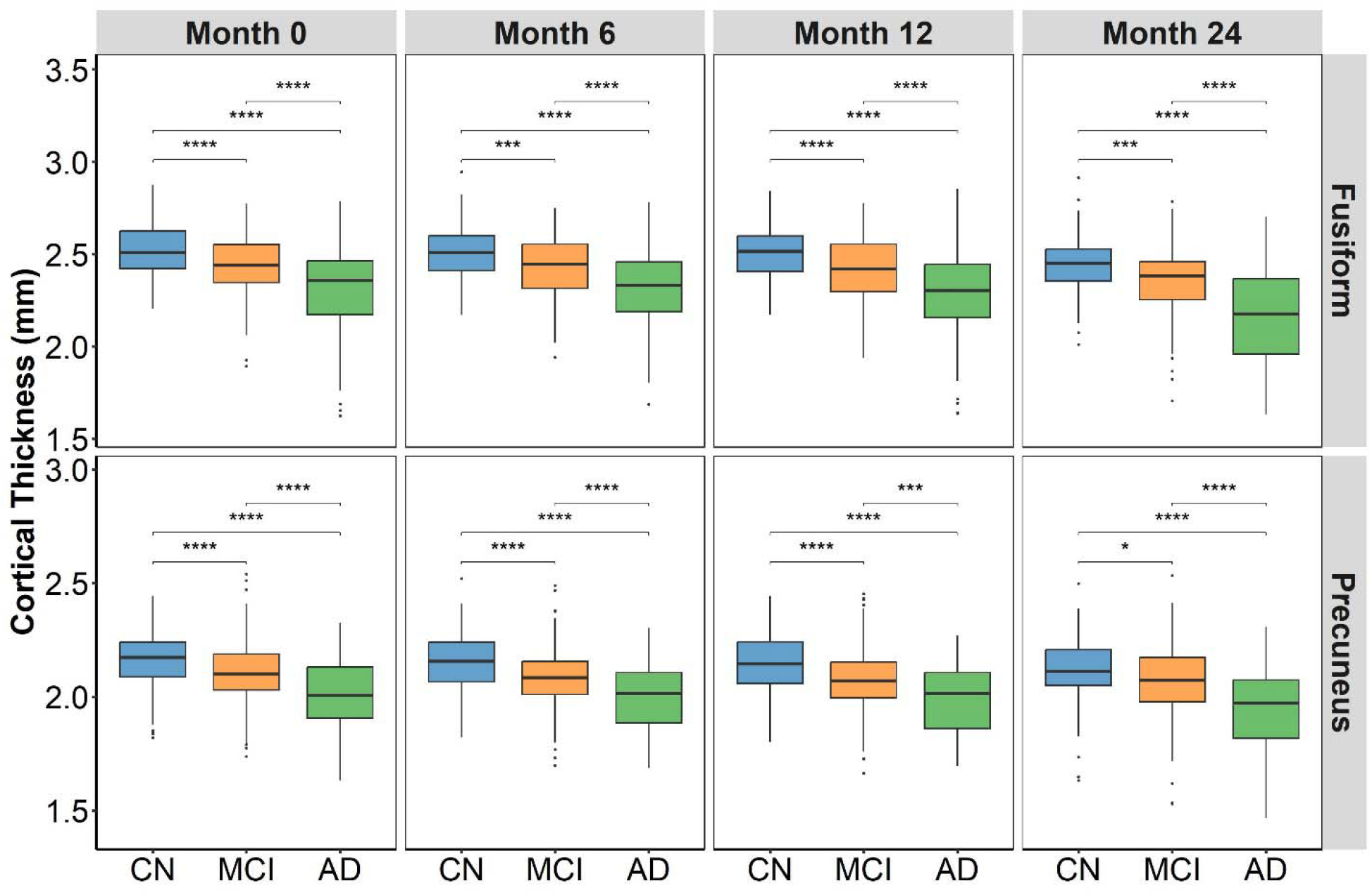
Boxplots illustrating CT of the fusiform gyrus and precuneus (right hemisphere) in CN, MCI, and AD individuals at 0-month, 6-month, 12-month and 24-month time point exhibiting progressive cortical thinning (*: p < 0.05; **: p < 0.01; ***: p < 0.001; ****: p < 0.0001).

**Figure 4:**
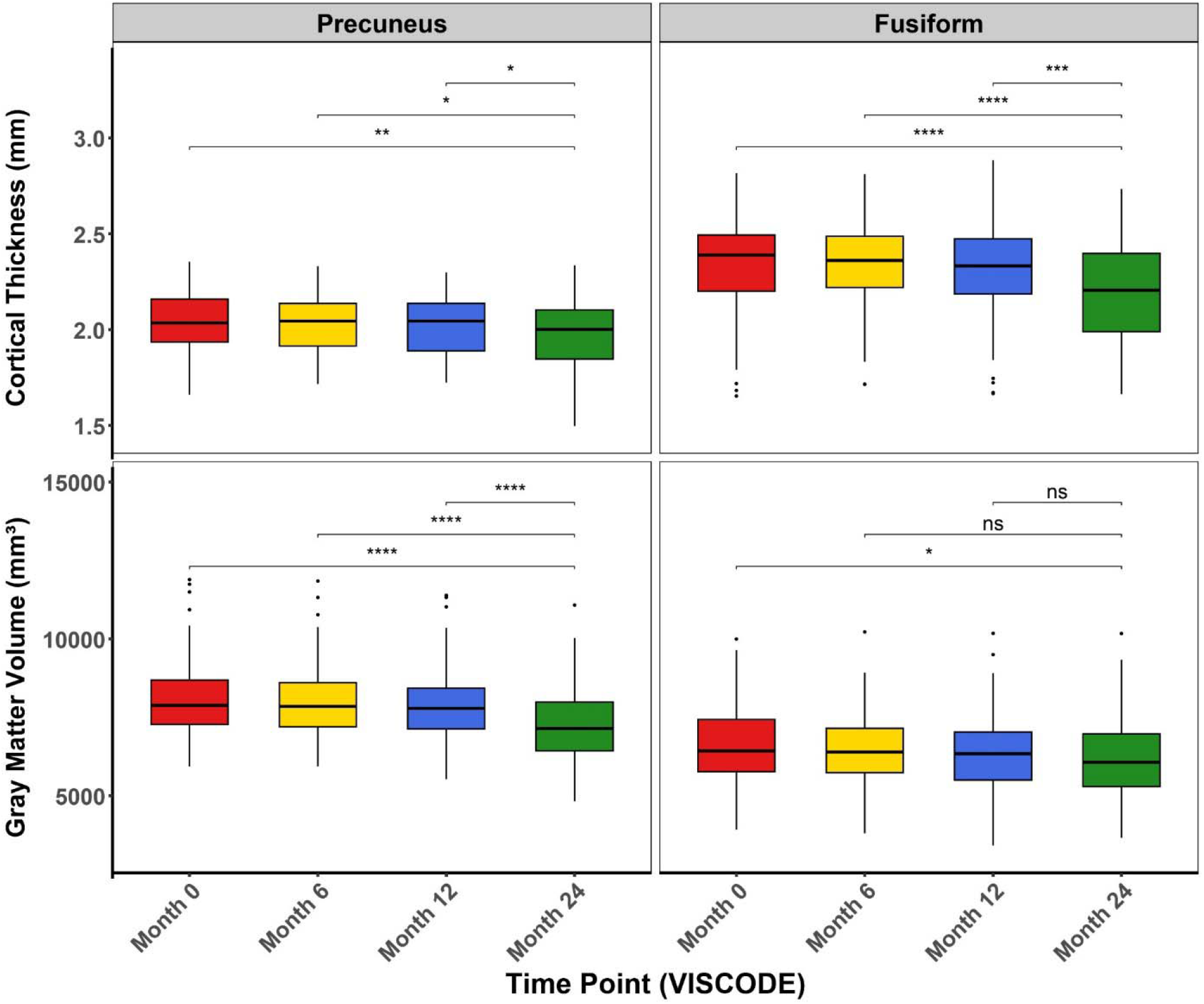
Boxplots illustrating GMV and CT of the fusiform gyrus and precuneus (right hemisphere) progression among AD individuals at 24-month time point (*: p < 0.05; **: p < 0.01; ***: p < 0.001; ****: p < 0.0001).

### Performance of Machine Learning-based Classification Models

We developed three classification models—AD vs CN, AD vs MCI, and MCI vs CN— using radiomic features from the precuneus and fusiform gyrus across both hemispheres. We first trained the models with age included as an additional feature. The AD vs CN model achieved an accuracy of 98.57%, AUC of 0.99, and MCC of 0.97 for the training dataset, and an accuracy of 85.71%, AUC of 0.94, and MCC of 0.71 for the testing dataset. Similarly, the AD vs MCI model showed an accuracy of 97.32%, AUC of 0.98, and MCC of 0.95 for the training dataset, and an accuracy of 81.70%, AUC of 0.90, and MCC of 0.62 for the testing dataset. The MCI vs CN model achieved an accuracy of 99.21%, AUC of 0.99, and MCC of 0.98 for the training dataset, and an accuracy of 87.40%, AUC of 0.93, and MCC of 0.75 for the testing dataset (Tables 1– 3, Figure 4)

**Table 1:** Performance of random forest classifier using radiomic features of both precuneus and fusiform gyrus (left and right hemisphere) for AD vs CN classification (n = Size of Dataset)

| **Dataset** |  | **Accuracy** | **Precision** | **Recall** | **F1-score** | **AUC** | **MCC** |
| --- | --- | --- | --- | --- | --- | --- | --- |
| **Without Age**<br>**( 36 features)** | **Training Dataset**<br>**(n = 839)** | 98.21% | 1.00 | 0.97 | 0.98 | 0.98 | 0.96 |
|  | **Testing Dataset**<br>**(n = 210)** | 84.29% | 0.86 | 0.87 | 0.86 | 0.93 | 0.68 |
| **With Age**<br>**( 37 features)** | **Training Dataset**<br>**(n = 839)** | 98.57% | 1.00 | 0.97 | 0.99 | 0.99 | 0.97 |
|  | **Testing Dataset**<br>**(n = 210)** | 85.71% | 0.88 | 0.88 | 0.88 | 0.94 | 0.71 |

**Table 2:**
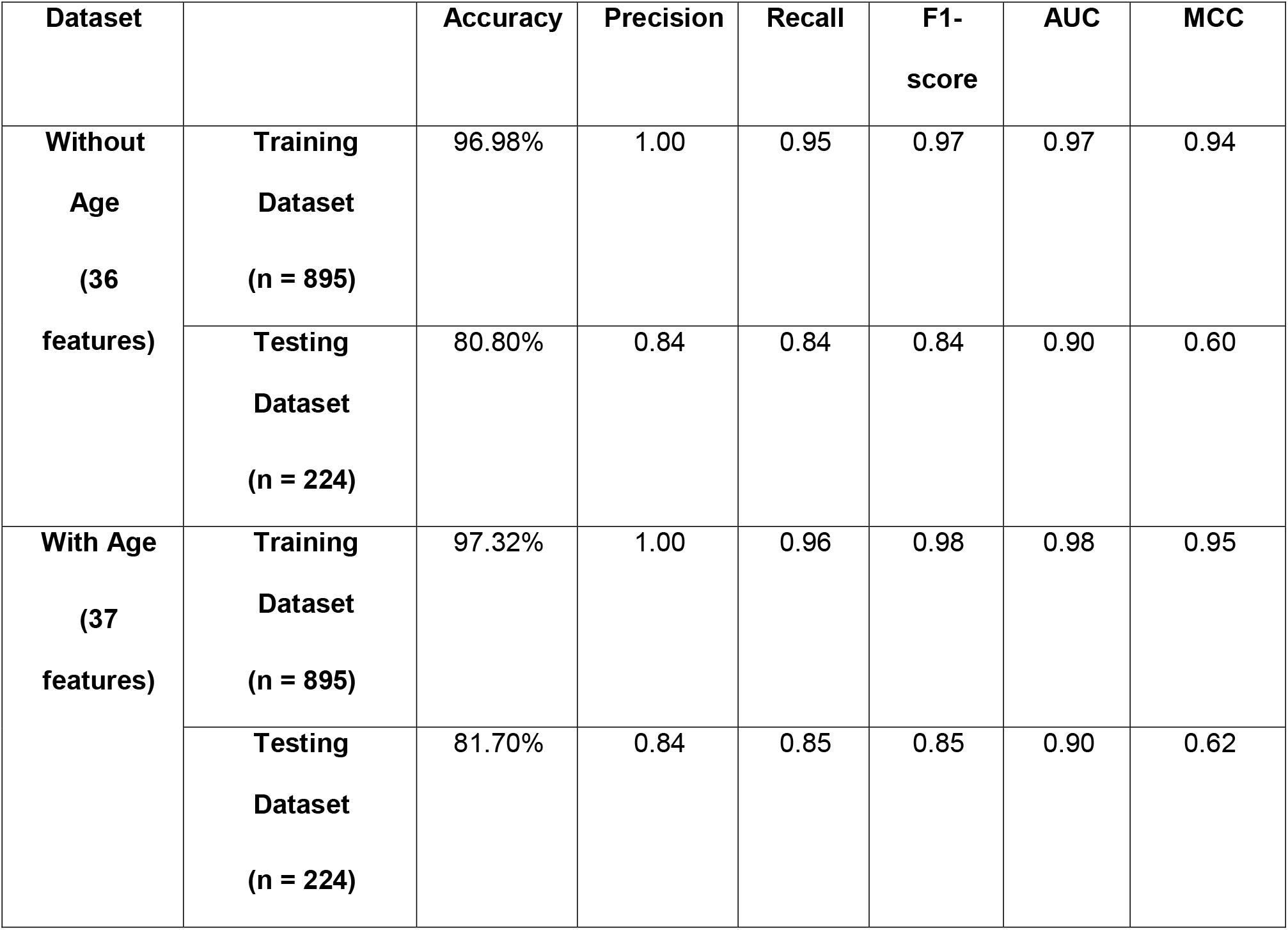
Performance of random forest classifier using radiomic features of both precuneus and fusiform gyrus (left and right hemisphere) for AD vs MCI classification (n = Size of Dataset).

**Table 3:** Performance of random forest classifier using radiomic features of both precuneus and fusiform gyrus (left and right hemisphere) for MCI vs CN classification (n = Size of Dataset).

| **Dataset** |  | **Accuracy** | **Precision** | **Recall** | **F1-score** | **AUC** | **MCC** |
| --- | --- | --- | --- | --- | --- | --- | --- |
| **Without Age (36 features)** | **Training Dataset (n = 1016)** | 99.31% | 1.00 | 0.99 | 0.99 | 0.99 | 0.99 |
|  | **Testing Dataset (n = 254)** | 84.25% | 0.84 | 0.87 | 0.85 | 0.90 | 0.68 |
| **With Age (37 features)** | **Training Dataset (n = 1016)** | 99.21% | 1.00 | 0.99 | 0.99 | 0.99 | 0.98 |
|  | **Testing Dataset (n = 254)** | 87.40 | 0.88 | 0.88 | 0.88 | 0.93 | 0.75 |

To mitigate the effect of age on model performance, we also trained the same models without including age as one of the training features. Interestingly, even without age, we observed similar performance across all three models. For the AD vs CN classifier, the model achieved an accuracy of 98.21%, AUC of 0.98, and MCC of 0.96 for the training dataset, and an accuracy of 84.29%, AUC of 0.93, and MCC of 0.68 for the testing dataset. For the AD vs MCI model, the training dataset showed an accuracy of 96.98%, AUC of 0.97, and MCC of 0.94, while the testing dataset showed an accuracy of 80.80%, AUC of 0.90, and MCC of 0.60. The MCI vs CN model achieved an accuracy of 99.31%, AUC of 0.99, and MCC of 0.99 for the training dataset, and an accuracy of 84.25%, AUC of 0.90, and MCC of 0.68 for the testing dataset (Figure 5). These results demonstrate that the inclusion of age led to only marginal gains in performance, reinforcing that the MRI-based radiomic features from the precuneus and fusiform gyrus are highly predictive on their own. Thus, the developed models demonstrate strong classification capability, independent of age, which has long been considered a strong determinant of AD

**Figure 5:**
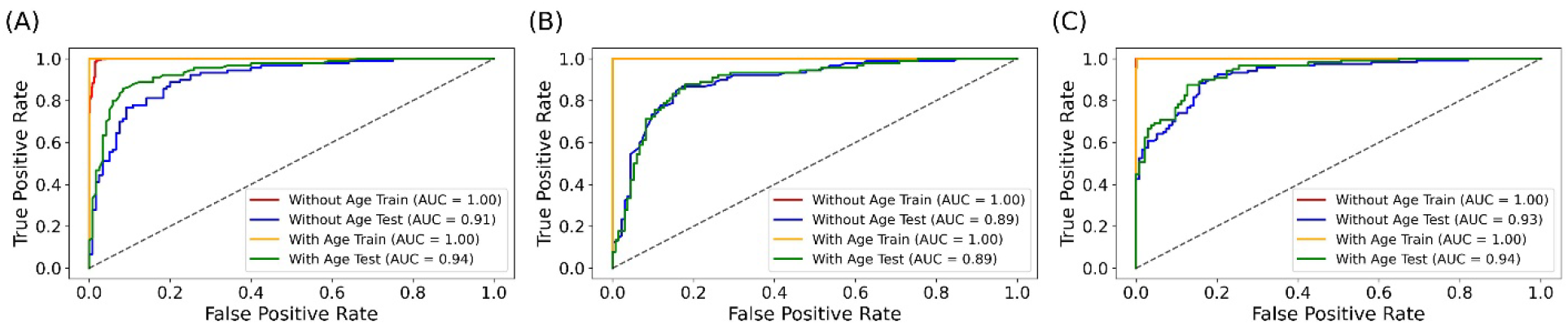
ROC plots for classification between (A) AD vs CN, (B) AD vs MCI, and (C) MCI vs CN a total of 36 radiomic features extracted from the fusiform gyrus and precuneus (18 from each region: 9 from the left and 9 from the right hemisphere). Classifier models were developed both with and without the inclusion of age.

To rigorously evaluate the independent discriminative power of each brain region, we trained separate classification models using radiomic features exclusively from the fusiform gyrus and the precuneus. These region-specific models demonstrated consistent and robust performance throughout. The classification models using fusiform gyrus-only radiomic features demonstrated strong performance across all diagnostic groups. For the AD vs CN classification with age included, an accuracy of 98.09%, AUC of 0.98, and MCC of 0.96 was obtained for the training dataset, and an accuracy of 82.86%, AUC of 0.91, and MCC of 0.65 for the testing dataset. Without age, an accuracy of 97.85%, AUC of 0.98, and MCC of 0.96 was achieved for the training dataset, and an accuracy of 82.86%, AUC of 0.90, and MCC of 0.65 for the testing dataset were achieved from the same model. For the AD vs MCI classification with age included, an accuracy of 96.87%, AUC of 0.97, and MCC of 0.94 was achieved for the training dataset, and an accuracy of 81.70%, AUC of 0.88, and MCC of 0.62 for the testing dataset. Without age, an accuracy of 96.20%, AUC of 0.97, and MCC of 0.92 was recorded for the training dataset, and an accuracy of 80.80%, AUC of 0.87, and MCC of 0.61 for the testing dataset. For the CN vs MCI classification with age included, an accuracy of 98.52%, AUC of 0.99, and MCC of 0.97 observed for the training dataset, and an accuracy of 79.74%, AUC of 0.86, and MCC of 0.57 for the testing dataset. Without age, an accuracy of 98.62%, AUC of 0.99, and MCC of 0.97 was achieved for the training dataset, and an accuracy of 73.62%, AUC of 0.82, and MCC of 0.47 for the testing dataset. (Supplementary Tables 2–4, Supplementary Figures 18).

The classification models using precuneus-only radiomic features also demonstrated consistent discriminative ability. For the AD VS CN classification with age included, an accuracy of 97.97%, AUC of 0.98, and MCC of 0.96 was recorded for the training dataset, and an accuracy of 83.81%, AUC of 0.92, and MCC of 0.67 for the testing dataset. Without age, 97.50%, AUC of 0.98, and MCC of 0.95 was achieved for the training dataset, and an accuracy of 81.90%, AUC of 0.88, and MCC of 0.63 for the testing dataset. For the AD vs MCI with age included, an accuracy of 96.98%, AUC of 0.97, and MCC of 0.94 was achieved for the training dataset, and an accuracy of 80.36%, AUC of 0.87, and MCC of 0.59 for the testing dataset. classification without age, an accuracy of 98.09%, AUC of 0.97, and MCC of 0.92 was obtained for the training dataset, and an accuracy of 79.46%, AUC of 0.86, and MCC of 0.57 for the testing dataset. For the CN vs MCI classification with age included, an accuracy of 98.82%, AUC of 0.99, and MCC of 0.98 was achieved for the training dataset, and an accuracy of 81.89%, AUC of 0.89, and MCC of 0.64 for the testing dataset. Without age, an accuracy of 98.52%, AUC of 0.99, and MCC of 0.97 was achieved for the training dataset, and an accuracy of 81.50%, AUC of 0.88, and MCC of 0.63 for the testing dataset. These results underscore the diagnostic relevance of both the fusiform gyrus and precuneus as standalone neuroimaging biomarkers. Their ability to support reliable classification between clinical stages of Alzheimer’s disease highlights their value for developing targeted, region-specific radiomic diagnostic models (Supplementary Tables 2–4, Supplementary Figures 19).

### Progression Prediction using Linear Regression Model and Time Series Model

For the right hemisphere features, the linear regression model yielded a correlation of 0.72 for fusiform GMV, with an RMSE of 873.49 and MAPE of 11.4. The time-series model improved upon this, yielding a correlation of 0.85, RMSE of 681.97, and MAPE of 8.82. For fusiform CT, the linear regression model achieved a correlation of 0.73, RMSE of 0.17, and MAPE of 6.71. The time-series model showed better performance with a correlation of 0.86, RMSE of 0.15, and MAPE of 5.59. The precuneus GMV produced a correlation of 0.80 in the linear regression model, along with an RMSE of 682.13 and MAPE of 8.26. The time-series model reached a higher correlation of 0.89, RMSE of 687.82, and MAPE of 8.2. For precuneus CT, the linear regression model achieved a correlation of 0.72, RMSE of 0.12, and MAPE of 5.15, while the time-series model improved these to a correlation of 0.84, RMSE of 0.10, and MAPE of 4.09.

For the left hemisphere features, the linear regression model demonstrated strong predictive performance. For fusiform GMV, it yielded a correlation of 0.97, RMSE of 319.42, and MAPE of 4.01, while the time-series model achieved a correlation of 0.98, RMSE of 323.7, and MAPE of 4.57. The fusiform Average Cortical Thickness showed a correlation of 0.93, RMSE of 0.09, and MAPE of 3.14 in the linear regression model, with improvements in the time-series model showing a correlation of 0.96, RMSE of 0.08, and MAPE of 3.1. For precuneus GMV, the linear regression model reached a correlation of 0.96, RMSE of 329.38, and MAPE of 3.98. The time-series model recorded a correlation of 0.97, RMSE of 299.74, and MAPE of 3.76. Lastly, precuneus CT demonstrated a correlation of 0.89, RMSE of 0.08, and MAPE of 3.38 using linear regression, while the time-series model further improved performance with a correlation of 0.94, RMSE of 0.07, and MAPE of 3.01. (Table 4). Overall, predictive performance was consistently higher for left hemisphere features compared to the right hemisphere across both modeling approaches (Supplementary Table 5).

**Table 4:** Performance of linear regression and time-series model using radiomic features of only AD to predict progression for GMV and CT.

| Features | Correlation (r) |  | MAPE |  | RMSE |  |
| --- | --- | --- | --- | --- | --- | --- |
|  | Linear Regression | Time Series | Linear Regression | Time Series | Linear Regression | Time Series |
| **RH Fusiform GMV** | 0.72 | 0.85 | 11.4 | 8.82 | 873.49 | 681.97 |
| **RH Fusiform CT** | 0.73 | 0.86 | 6.71 | 5.59 | 0.17 | 0.15 |
| **RH Precuneus GMV** | 0.8 | 0.89 | 8.26 | 8.2 | 682.13 | 687.82 |
| **RH Precuneus CT** | 0.72 | 0.84 | 5.15 | 4.09 | 0.12 | 0.1 |
| **LH Fusiform GMV** | 0.97 | 0.98 | 4.01 | 4.57 | 319.42 | 323.7 |
| **LH Fusiform CT** | 0.93 | 0.96 | 3.14 | 3.1 | 0.09 | 0.08 |
| **LH Precuneus GMV** | 0.96 | 0.97 | 3.98 | 3.76 | 329.38 | 299.74 |
| **LH Precuneus CT** | 0.89 | 0.94 | 3.38 | 3.01 | 0.08 | 0.07 |

RH: Right Hemisphere; LH: Left Hemisphere; GMV: Grey Matter Volume; CT: Cortical Thickness; MAPE: Mean Absolute Percentage Error; RMSE: Root Mean Square Error.

## Discussion

This study explored potential structural associations between the precuneus and fusiform gyrus with AD pathology using statistical and machine learning analysis on MRI-based radiomic features. We extracted nine MRI-based radiomic features of the left and right hemispheres of the precuneus and fusiform gyrus i.e., GMV, CT, folding index, intrinsic curvature index, integrated rectified Gaussian curvature, integrated rectified mean curvature, surface area, and number of vertices using FreeSurfer automated pipeline. In our statistical analysis, we observed that GMV of both precuneus and fusiform significantly decreased in AD patients as compared to the MCI and CN in follow up data of four time points (0, 6, 12 and 24 months). Reduced GMV is a prevalent sign of AD pathology and has been proposed as an essential physiological structure of cognitive function decline (31). Our observation aligns with a previous report on Voxel-based research that found GMV reductions in the right fusiform gyrus of AD patients (32). Moreover, lower gray matter density in the precuneus and its atrophy was shown to be more prominent in early-onset AD patients in various voxel-based investigations on gray matter atrophy (15, 33). Additionally, we also observed a significant decrease in average thickness of precuneus and fusiform gyrus in AD patients. There was significant increase in gaussian curvature of AD patients in fusiform gyrus. Significant reduction in CT was previously reported in patients with AD and MCI (32), however increased gaussian curvature along with a decrease in precuneus and fusiform thickness were not reported earlier. Our findings reinforce the involvement of the precuneus and fusiform gyrus in AD pathology and offer new insights into their potential for early detection and diagnosis

To further investigate the potential of the precuneus and fusiform in classifying AD patients with MCI and CN, we developed random forest-based classifier models. we developed three binary classification models using radiomic features extracted from the precuneus and fusiform gyrus (37 features in total, including age). The models demonstrated strong discriminative performance, training accuracies of 98.57% for AD vs. CN, 97.32% for AD vs. MCI, and 99.21% for MCI vs. CN. To mitigate the effect of age on model performance, we also constructed models excluding age as a feature. Interestingly, removing age as a training feature had negligible effect on model performance (98.21% for AD vs. CN, 96.98% for AD vs. MCI, and 99.31% for MCI vs. CN). These results suggest that precuneus and fusiform gyrus based radiomic features have robust diagnostic potential, even without age.

Given the availability of longitudinal data across multiple follow-up time points, we further investigated the progression of AD using both linear regression and time-series models. Our linear regression and time-series model analysis provides additional evidence for the utility of radiomic features of the precuneus and fusiform gyrus in predicting AD progression over time. The results demonstrate that radiomic features from both hemispheres are effective in predicting AD progression, with the time-series model consistently outperforming linear regression. Across GMV and CT features, the time-series model achieved higher correlation values (r = 0.84–0.98) and lower MAPE (3.01–8.82) compared to the linear regression model (r = 0.72–0.97, MAPE 3.14–11.4). Left hemisphere features generally exhibited stronger predictive performance than those from the right hemisphere. These findings highlight the advantage of temporal modeling and the predictive strength of structural brain features in tracking AD progression.

We achieved high performances in both classification and regression models, which suggests their potential clinical applicability of these models and radiomic features as biomarkers for early detection and monitoring disease progression. Our findings align with previous literature emphasizing the structural importance of these brain regions in AD pathology. Further investigations are necessary to expand the knowledge and understanding of the precise pathways through which the precuneus and fusiform gyrus contribute to the AD pathology. A better understanding of the role of the precuneus and fusiform gyrus and feature-based analysis in AD individuals could provide a generalized early detection method.

## Conclusion

To evaluate the diagnostic and prognostic potential of these radiomic features, we developed a random forest classifier and assessed its performance using Accuracy, Area Under the Curve (AUC), and Matthews Correlation Coefficient (MCC). For modeling disease progression, we compared a linear regression model and a time-series model, using standard evaluation metrics including Pearson correlation coefficient (r), Mean Absolute Percentage Error (MAPE), and Root Mean Square Error (RMSE). Results from both classification and regression approaches underscore the reliability and utility of radiomic biomarkers from the precuneus and fusiform gyrus.Collectively, these findings highlight the promise of radiomic features as non-invasive biomarkers for early detection, longitudinal monitoring, and stratification of AD progression. Future work should focus on validating these findings in larger, independent cohorts and integrating radiomic data with cognitive assessments, functional neuroimaging, and genomic profiles to advance multi-modal frameworks for AD diagnosis and personalized intervention.

In conclusion, this study is among the first to comprehensively demonstrate the combined contribution of the precuneus and fusiform gyrus in AD pathology using radiomics. These insights enhance our understanding of the neuroanatomical basis of

AD and offer a foundation for more targeted and timelier diagnostic and therapeutic strategies.

## Abbreviations

AD: Alzheimer’s Disease
MCI: Mild Cognitive Impairment
CN: Cognitively Normal
MRI: Magnetic Resonance Imaging
GMV: Gray Matter Volume
CT: Cortical Thickness
ADNI: Alzheimer’s Disease Neuroimaging Initiative
MMSE: Mini-Mental State Examination
CDR: Clinical Dementia Rating
NINCDS/ADRDA: National Institute of Neurological and Communicative Disorders and Stroke / Alzheimer’s Disease and Related Disorders Association
FOV: Field of View
TR: Repetition Time
RFC: Random Forest Classifier
KNN: K-Nearest Neighbors
ROC AUC: Receiver Operating Characteristic – Area Under the Curve
MCC: Matthews Correlation Coefficient
MAE: Mean Absolute Error
RMSE: Root Mean Square Error
R^2^: Coefficient of Determination (R-squared)

## Acknowledgements

Authors acknowledge the research infrastructure provided by the Indian Institute of Technology Hyderabad.

## Author Contribution

Conceptualization: ISK, SS, NK, and RK; Investigation: ISK, SS, KK, SSK, RKh, AM, NK, and RK; Methodology: ISK, SS, KK, SSK, RKh, AM, NK, and RK; Project administration: ISK, NK, and RK; Software: ISK, SS, and RKh; Writing-original draft: ISK, SS, KK, SSK, RKh, AM, NK, and RK; Writing-review and editing: ISK, SS, RKh, AM, NK, and RK. All authors read and approved the final manuscript

## Data Availability

MRI scans were from Alzheimer’s Disease Neuroimaging Initiative (ADNI) database (http://adni.loni.usc.edu/).

## Code availability

All the models generated in this study are available on the following GitHub repository: (https://github.com/CGnTLab/AD_Radiomics).

