## Supplementary Figures for "Evaluation of Precuneus and Fusiform Gyrus-Based Radiomic biomarkers for Alzheimer’s disease Classification and Progression"

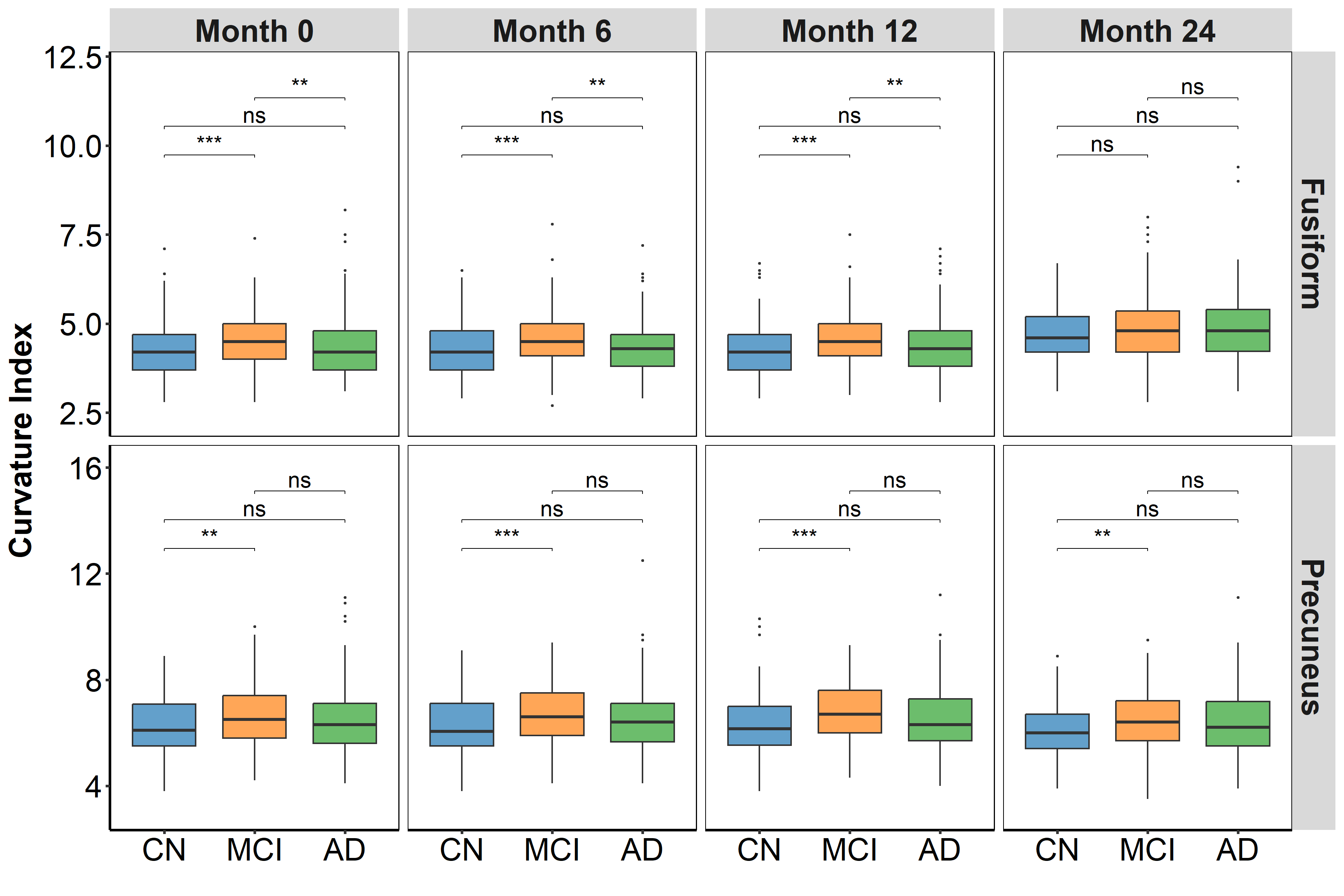

**Supp. Figure 1:** Curvature index (CI) of representative samples from cognitive normal (CN), mild cognitive impairment (MCI), and Alzheimer's disease (AD) individuals at three separate time points (months 0, 6, and 12). A statistically significant difference (*: p ≤ 0.05; **: p ≤ 0.01; ***: p ≤ 0.001; ****: p ≤ 0.0001; ns: p > 0.05) is shown in a box plot based on the Mann-Whitney U test for CI of the right hemisphere of precuneus and fusiform gyrus.

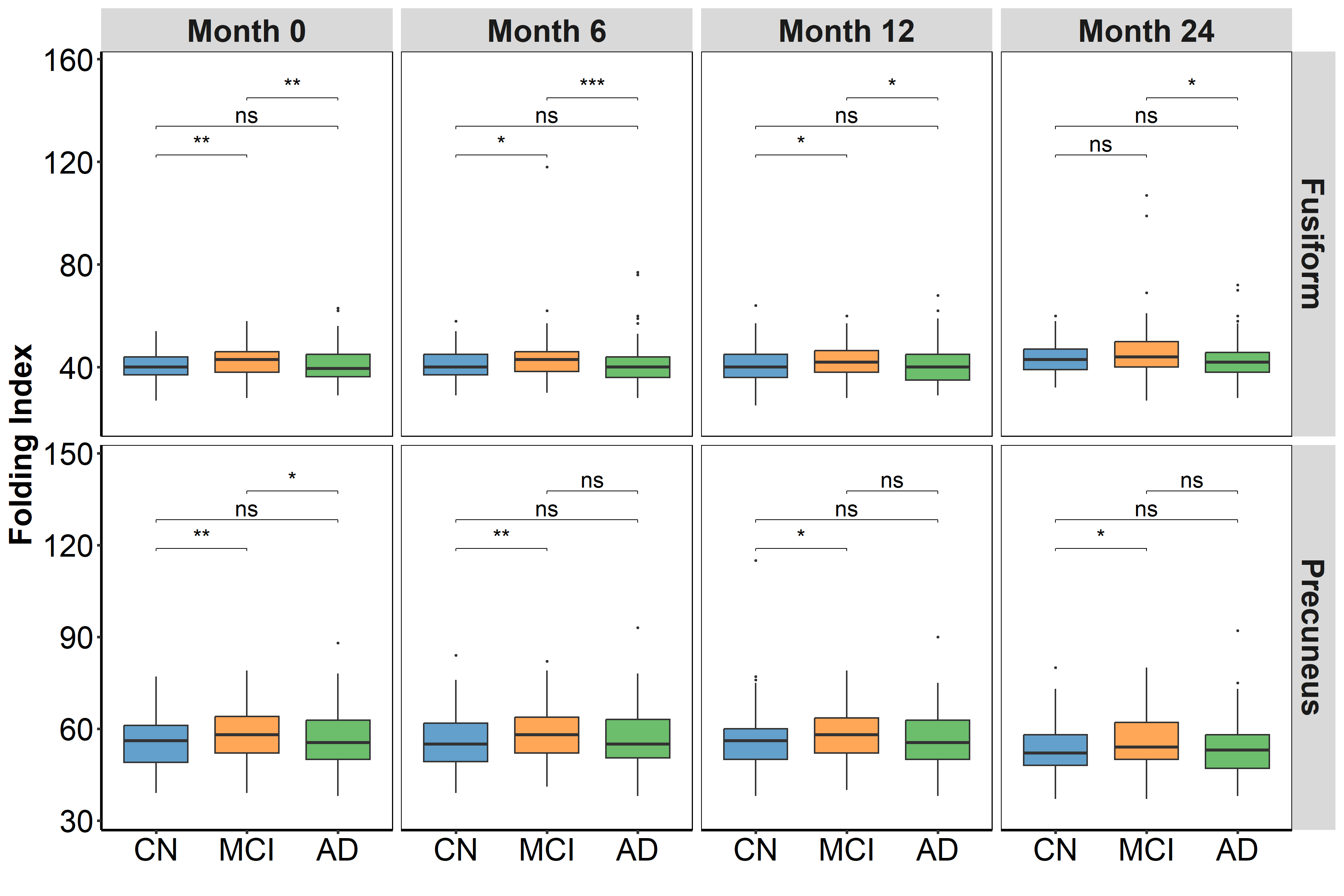

**Supp. Figure 2:** Folding index (FI) of representative samples from cognitive normal (CN), mild cognitive impairment (MCI), and Alzheimer's disease (AD) individuals at three separate time points (months 0, 6, and 12). A statistically significant difference (*: p ≤ 0.05; **: p ≤ 0.01; ***: p ≤ 0.001; ****: p ≤ 0.0001; ns: p > 0.05) is shown in a box plot based on the Mann-Whitney U test for FI of the right hemisphere of precuneus and fusiform gyrus.

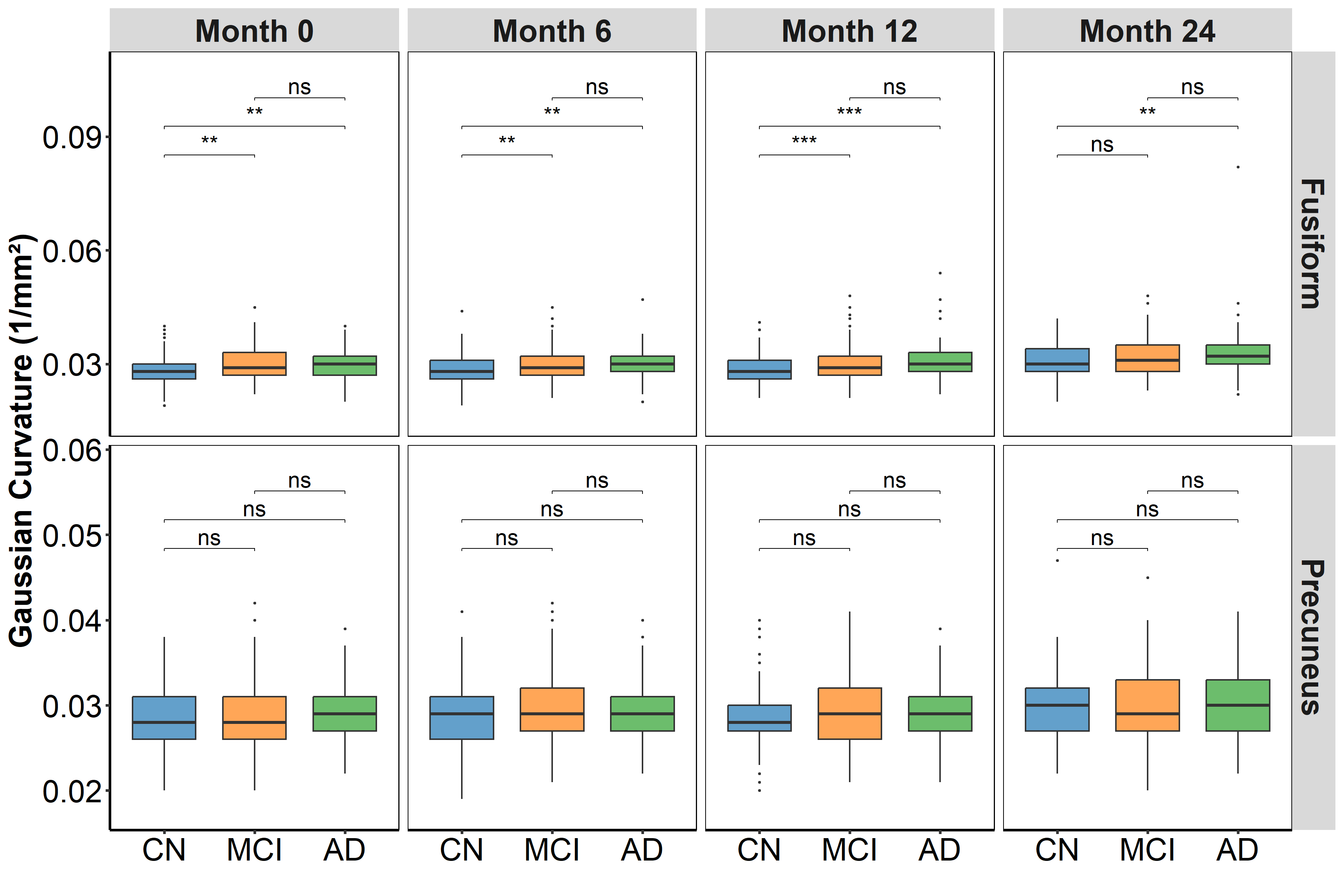

**Supp. Figure 3:** Gaussian curvature (GC) of representative samples from cognitive normal (CN), mild cognitive impairment (MCI), and Alzheimer's disease (AD) individuals at three different time points (months 0, 6, and 12). A statistically significant difference (*: p ≤ 0.05; **: p ≤ 0.01; ***: p ≤ 0.001; ****: p ≤ 0.0001; ns: p > 0.05) is shown in a box plot based on the Mann-Whitney U test for GC of the right hemisphere of precuneus and fusiform gyrus

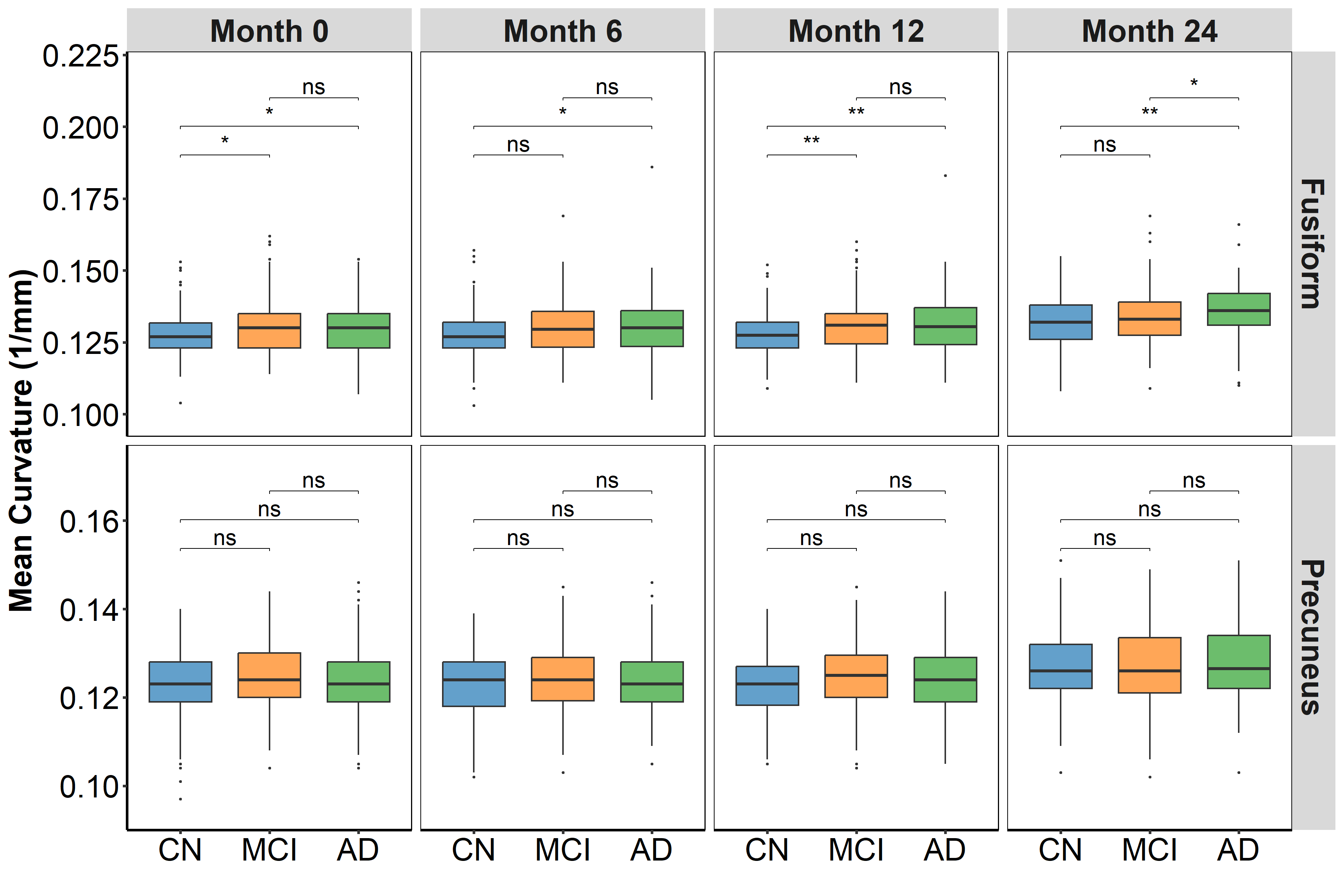

**Supp. Figure 4:** Mean curvature (MC) of representative samples from cognitive normal (CN), mild cognitive impairment (MCI), and Alzheimer's disease (AD) individuals at three different time points (months 0, 6, and 12). A statistically significant difference (*: p ≤ 0.05; **: p ≤ 0.01; ***: p ≤ 0.001; ****: p ≤ 0.0001; ns: p > 0.05) is shown in a box plot based on the Mann-Whitney U test for MC of the right hemisphere of precuneus and fusiform gyrus.

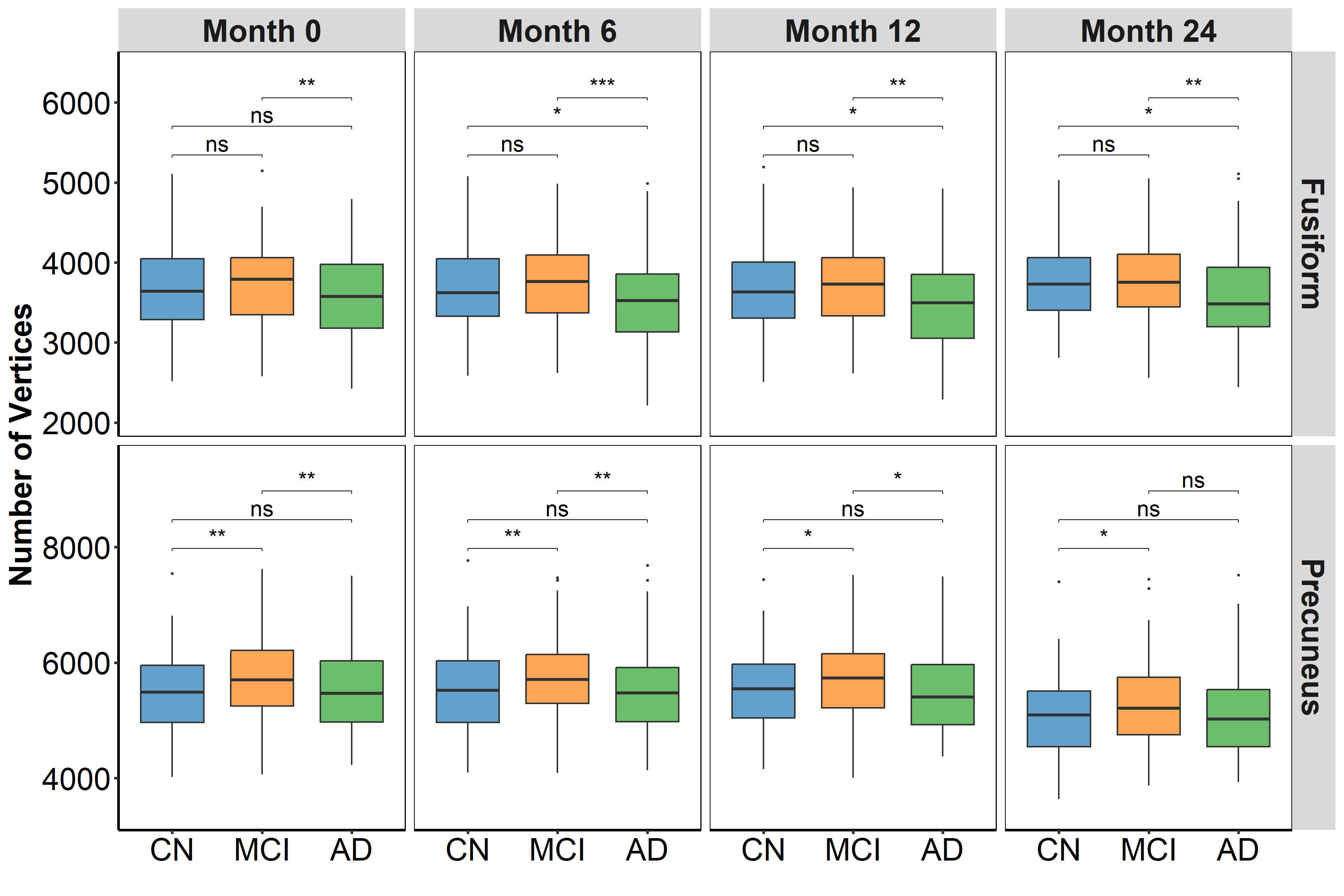

**Supp. Figure 5:** Number of vertices (NV) of representative samples from cognitive normal (CN), mild cognitive impairment (MCI), and Alzheimer's disease (AD) individuals at three separate time points (months 0, 6, and 12). A statistically significant difference (*: p ≤ 0.05; **: p ≤ 0.01; ***: p ≤ 0.001; ****: p ≤ 0.0001; ns: p > 0.05) is shown in a box plot based on the Mann-Whitney U test for NV of the right hemisphere of precuneus and fusiform gyrus.

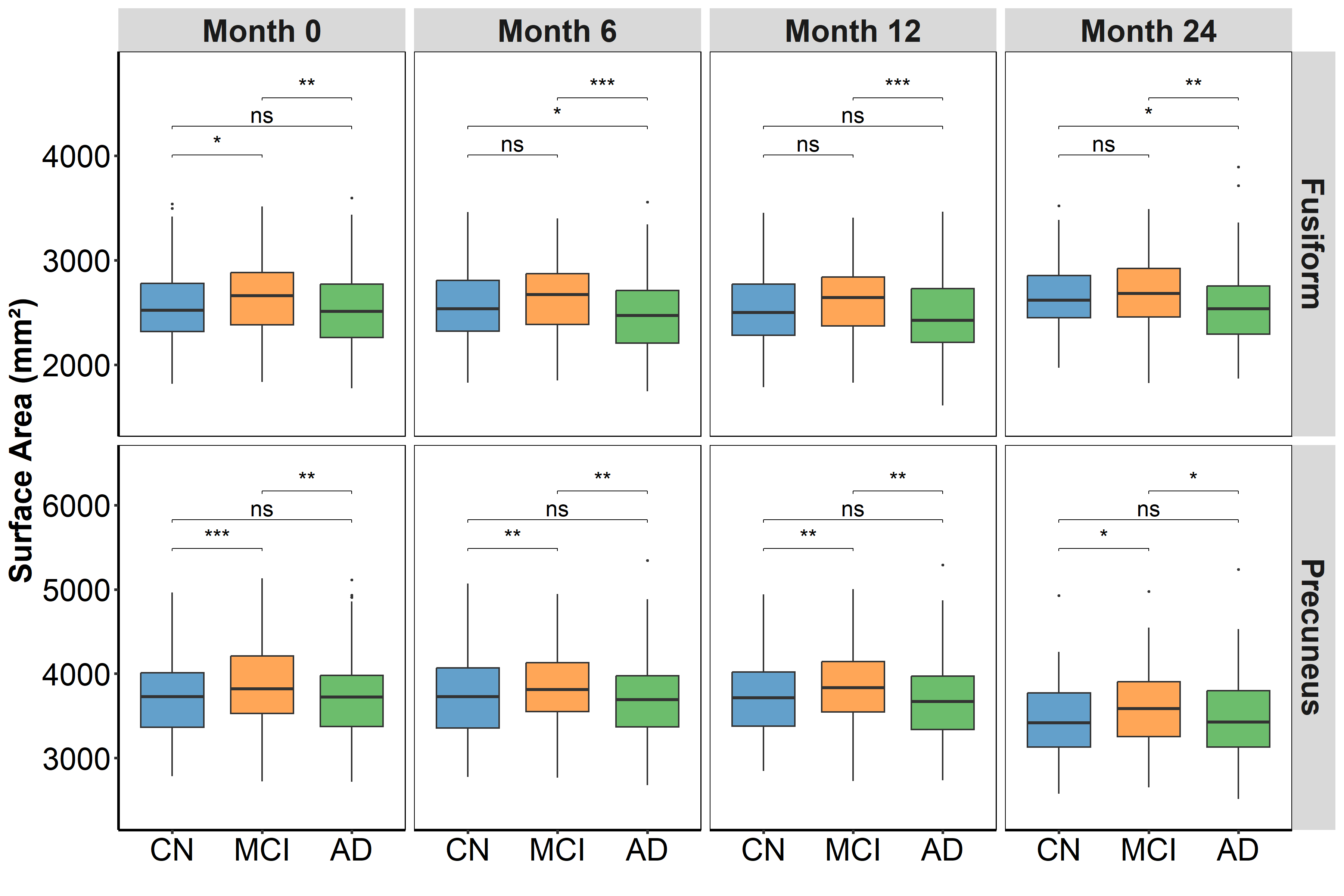

**Supp. Figure 6:** Surface area (SA) of representative samples from cognitive normal (CN), mild cognitive impairment (MCI), and Alzheimer's disease (AD) individuals at three separate time points (months 0, 6, and 12). A statistically significant difference (*: p ≤ 0.05; **: p ≤ 0.01; ***: p ≤ 0.001; ****: p ≤ 0.0001; ns: p > 0.05) is shown in a box plot based on the Mann-Whitney U test for SA of the right hemisphere of precuneus and fusiform gyrus.

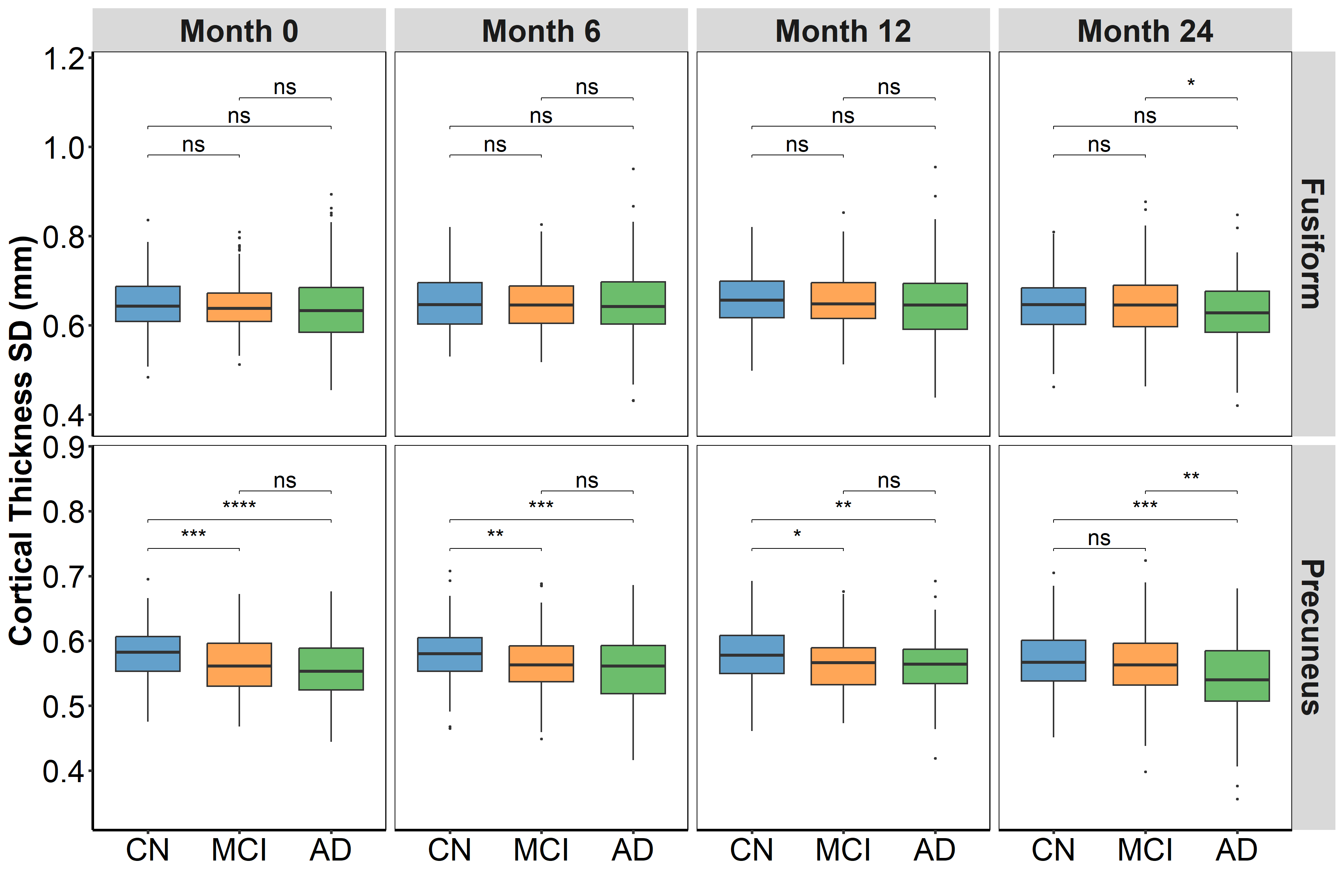

**Supp. Figure 7:** Cortical thickness Standard (CTS) of representative samples from cognitive normal (CN), mild cognitive impairment (MCI), and Alzheimer's disease (AD) individuals at three different time points (months 0, 6, and 12). A statistically significant difference (*: p ≤ 0.05; **: p ≤ 0.01; ***: p ≤ 0.001; ****: p ≤ 0.0001) is shown in a box plot based on the Mann-Whitney U test for CT of the right hemisphere of precuneus and fusiform gyrus.

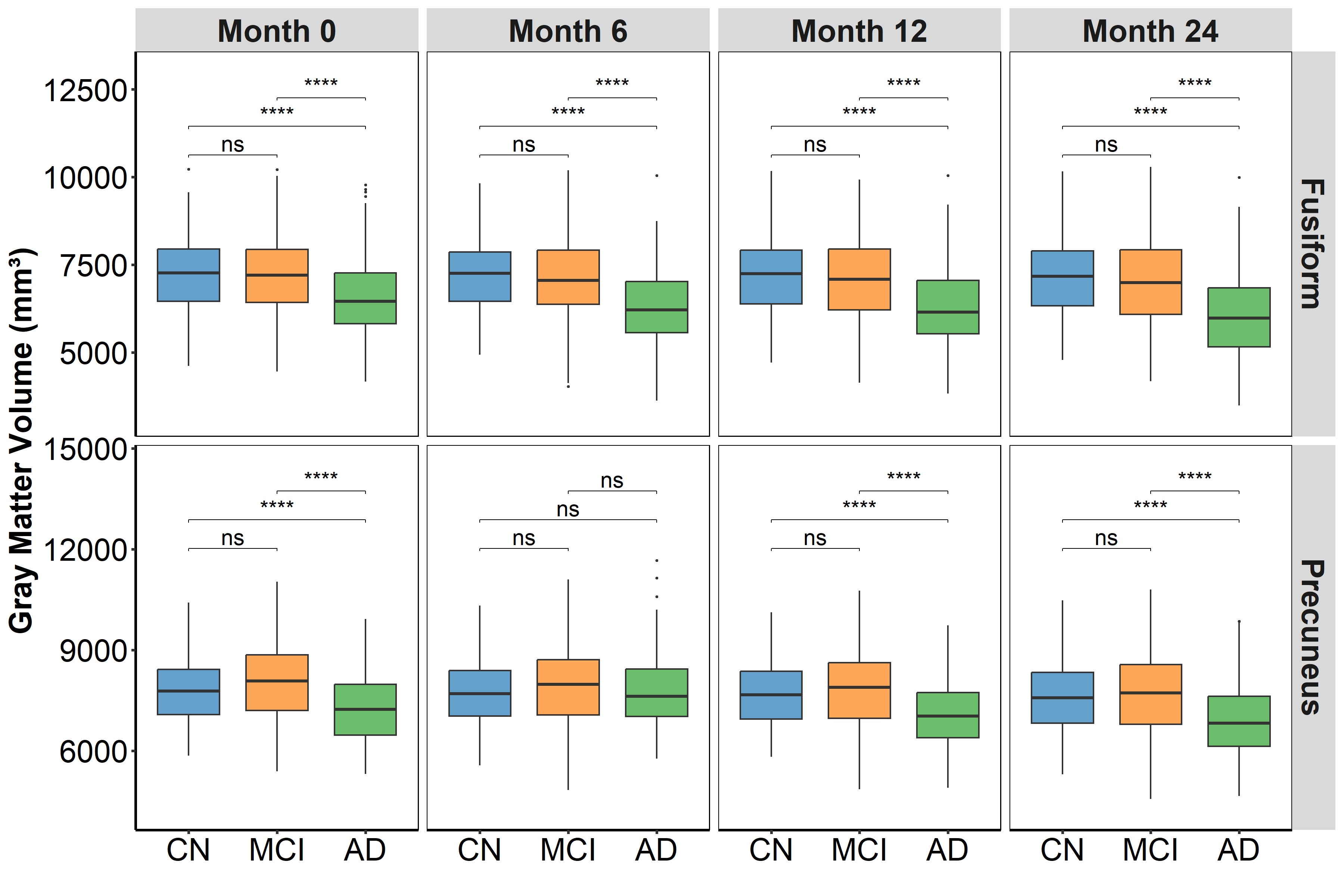

**Supp. Figure 8:** Grey Matter Volume (GMV) of representative samples from cognitive normal (CN), mild cognitive impairment (MCI), and Alzheimer's disease (AD) individuals at three different time points (months 0, 6, and 12). A statistically significant difference (*: p ≤ 0.05; **: p ≤ 0.01; ***: p ≤ 0.001; ****: p ≤ 0.0001) is shown in a box plot based on the Mann-Whitney U test for CT of the left hemisphere of precuneus and fusiform gyrus.

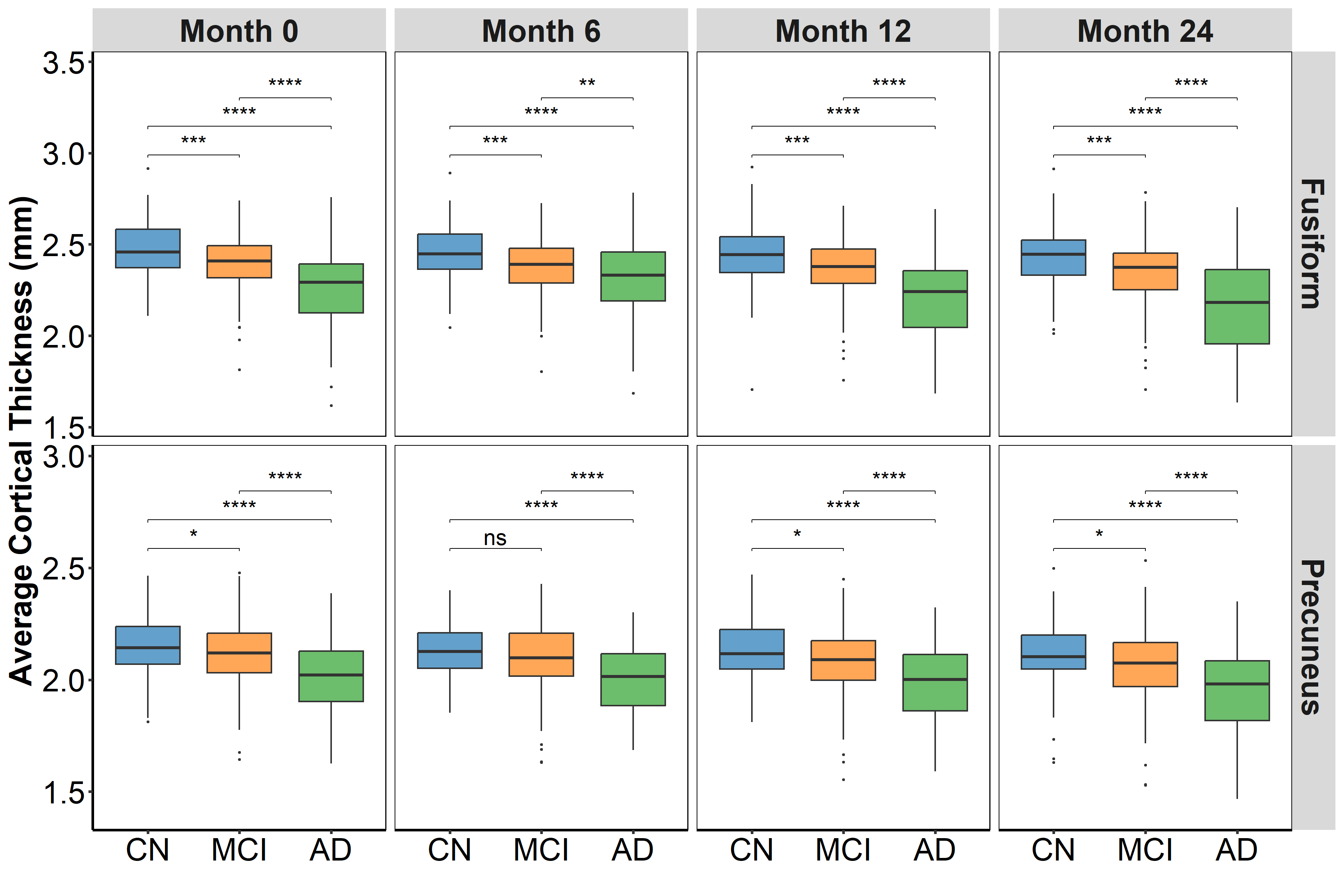

**Supp. Figure 9:** Cortical Thickness of representative samples from cognitive normal (CN), mild cognitive impairment (MCI), and Alzheimer's disease (AD) individuals at three different time points (months 0, 6, and 12). A statistically significant difference (*: p ≤ 0.05; **: p ≤ 0.01; ***: p ≤ 0.001; ****: p ≤ 0.0001) is shown in a box plot based on the Mann-Whitney U test for CT of the left hemisphere of precuneus and fusiform gyrus.

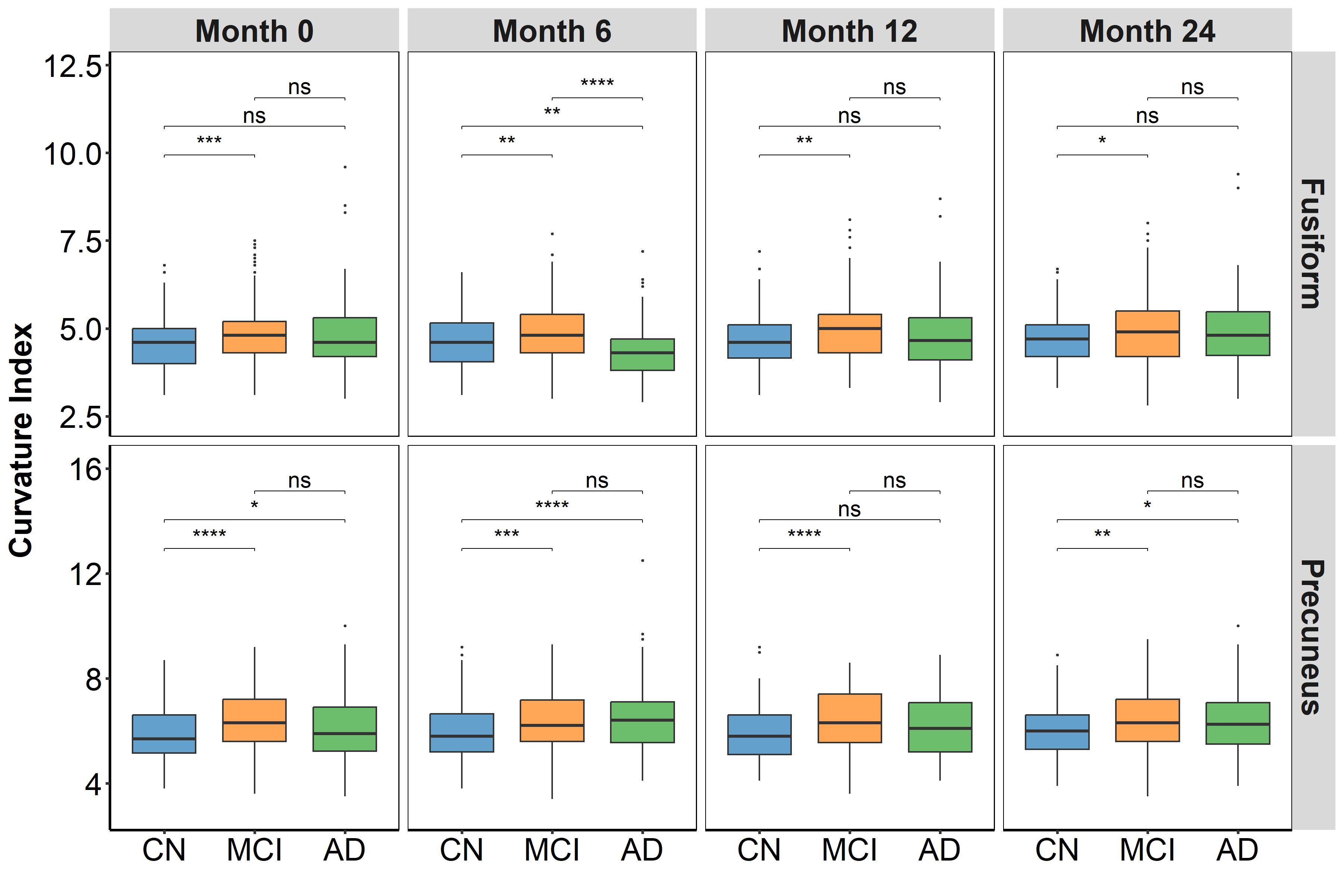

**Supp. Figure 10 :** Curvature index (CI) of representative samples from cognitive normal (CN), mild cognitive impairment (MCI), and Alzheimer's disease (AD) individuals at three separate time points (months 0, 6, and 12). A statistically significant difference (*: p ≤ 0.05; **: p ≤ 0.01; ***: p ≤ 0.001; ****: p ≤ 0.0001; ns: p > 0.05) is shown in a box plot based on the Mann-Whitney U test for CI of the left hemisphere of precuneus and fusiform gyrus.

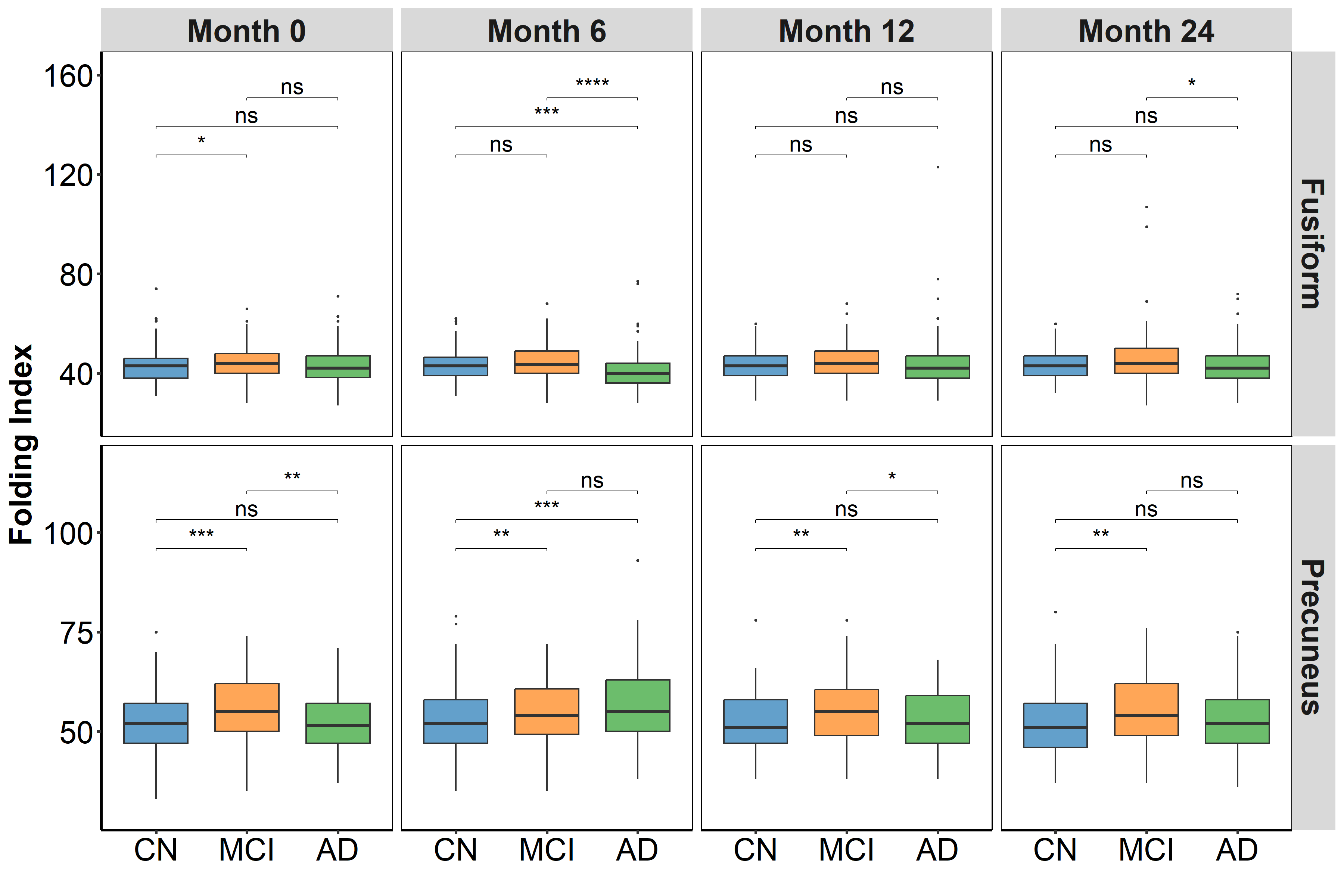

**Supp. Figure 11:** Folding index (FI) of representative samples from cognitive normal (CN), mild cognitive impairment (MCI), and Alzheimer's disease (AD) individuals at three separate time points (months 0, 6, and 12). A statistically significant difference (*: p ≤ 0.05; **: p ≤ 0.01; ***: p ≤ 0.001; ****: p ≤ 0.0001; ns: p > 0.05) is shown in a box plot based on the Mann-Whitney U test for FI of the left hemisphere of precuneus and fusiform gyrus.

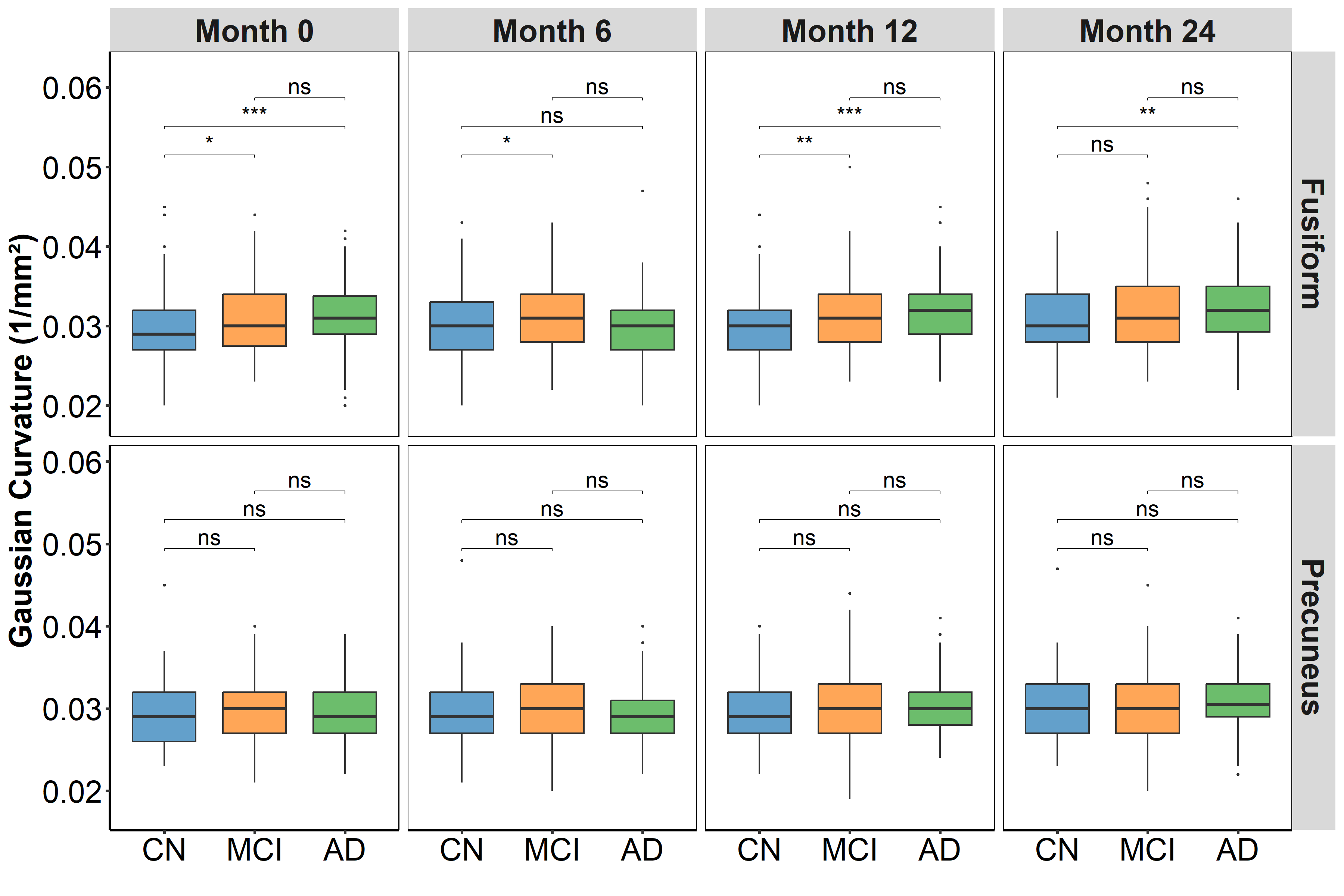

**Supp. Figure 12:** Gaussian curvature (GC) of representative samples from cognitive normal (CN), mild cognitive impairment (MCI), and Alzheimer's disease (AD) individuals at three different time points (months 0, 6, and 12). A statistically significant difference (*: p ≤ 0.05; **: p ≤ 0.01; ***: p ≤ 0.001; ****: p ≤ 0.0001; ns: p > 0.05) is shown in a box plot based on the Mann-Whitney U test for GC of the left hemisphere of precuneus and fusiform gyrus

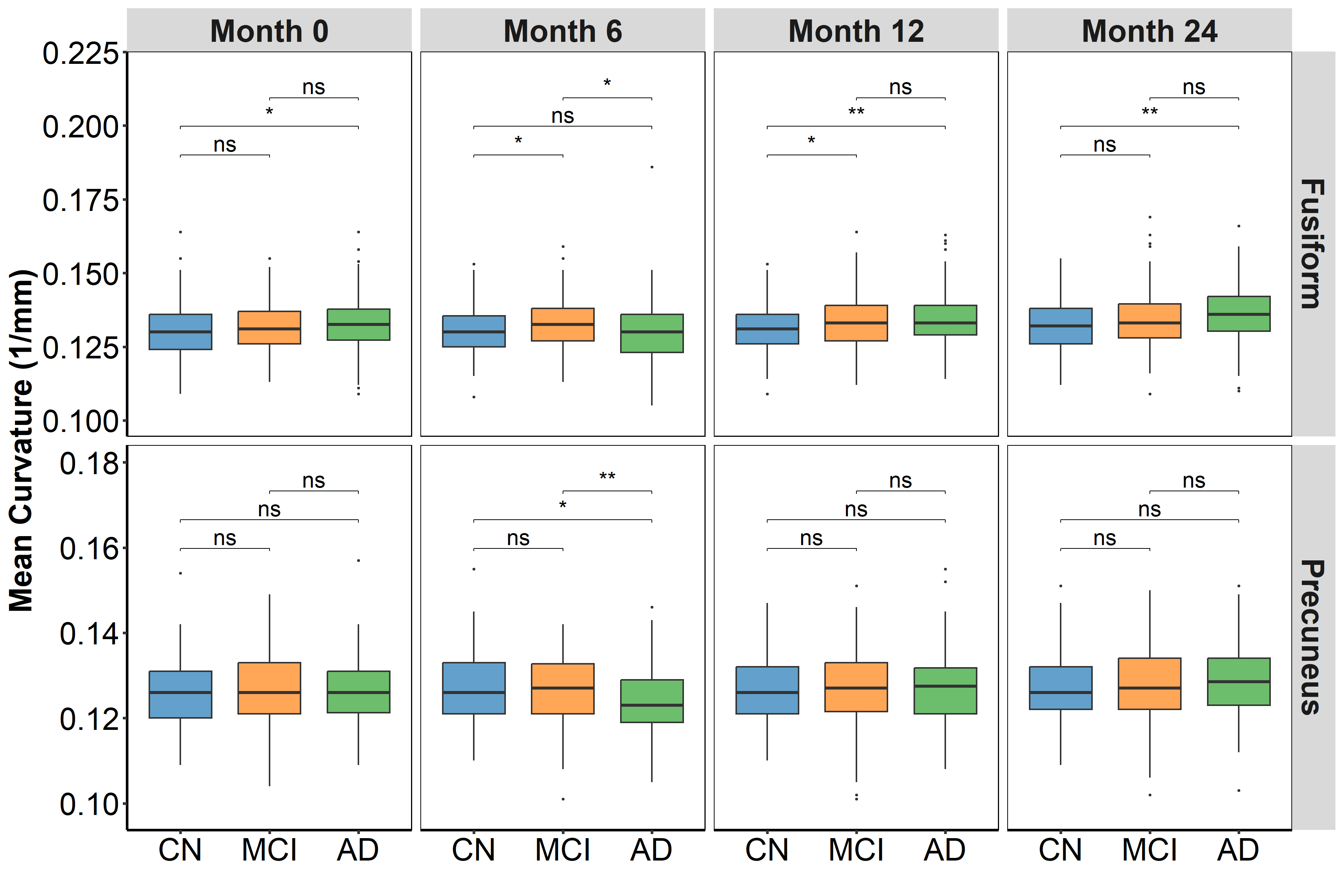

**Supp. Figure 13:** Mean curvature (MC) of representative samples from cognitive normal (CN), mild cognitive impairment (MCI), and Alzheimer's disease (AD) individuals at three different time points (months 0, 6, and 12). A statistically significant difference (*: p ≤ 0.05; **: p ≤ 0.01; ***: p ≤ 0.001; ****: p ≤ 0.0001; ns: p > 0.05) is shown in a box plot based on the Mann-Whitney U test for MC of the left hemisphere of precuneus and fusiform gyrus.

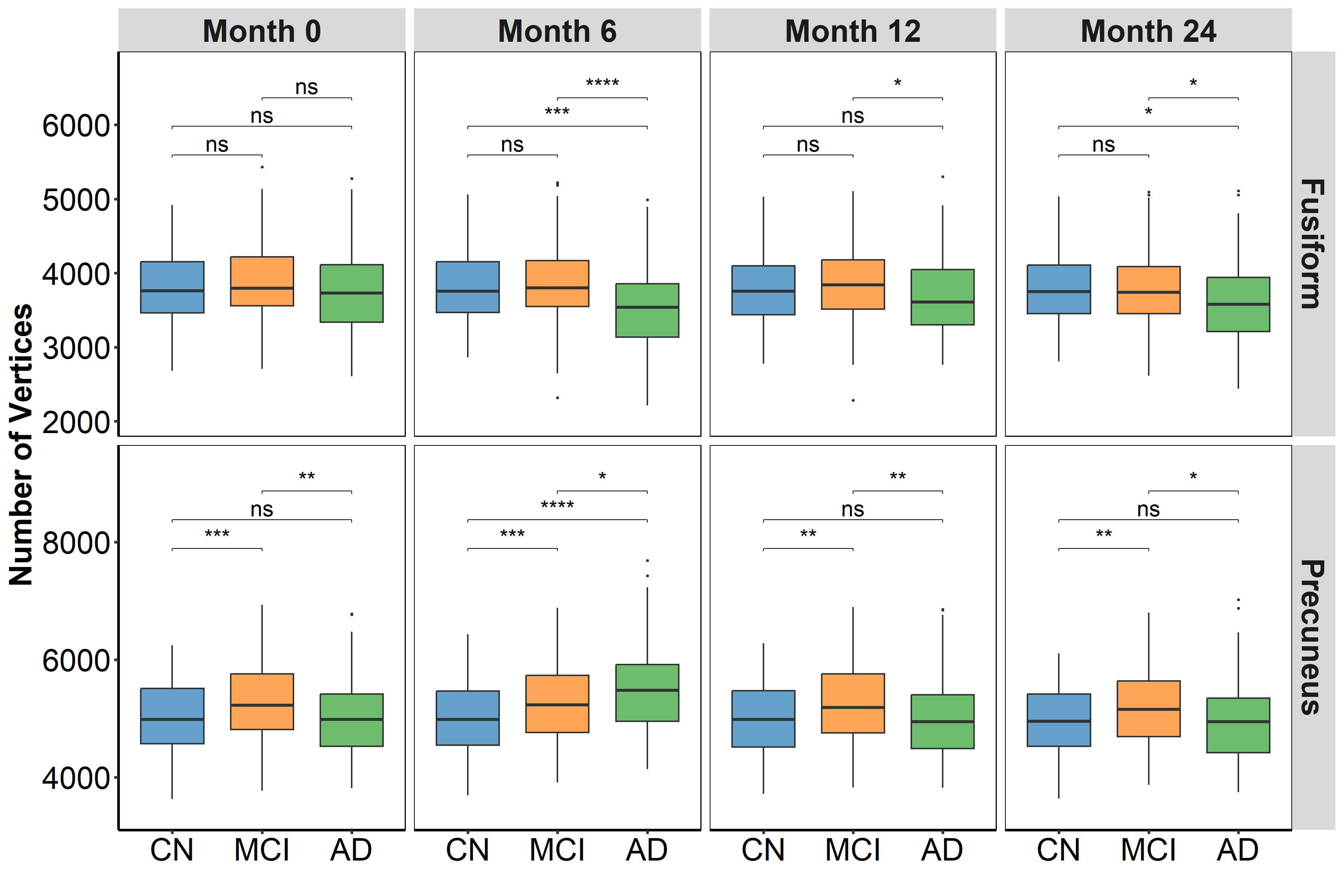

**Supp. Figure 14:** Number of vertices (NV) of representative samples from cognitive normal (CN), mild cognitive impairment (MCI), and Alzheimer's disease (AD) individuals at three separate time points (months 0, 6, and 12). A statistically significant difference (*: p ≤ 0.05; **: p ≤ 0.01; ***: p ≤ 0.001; ****: p ≤ 0.0001; ns: p > 0.05) is shown in a box plot based on the Mann-Whitney U test for NV of the left hemisphere of precuneus and fusiform gyrus.

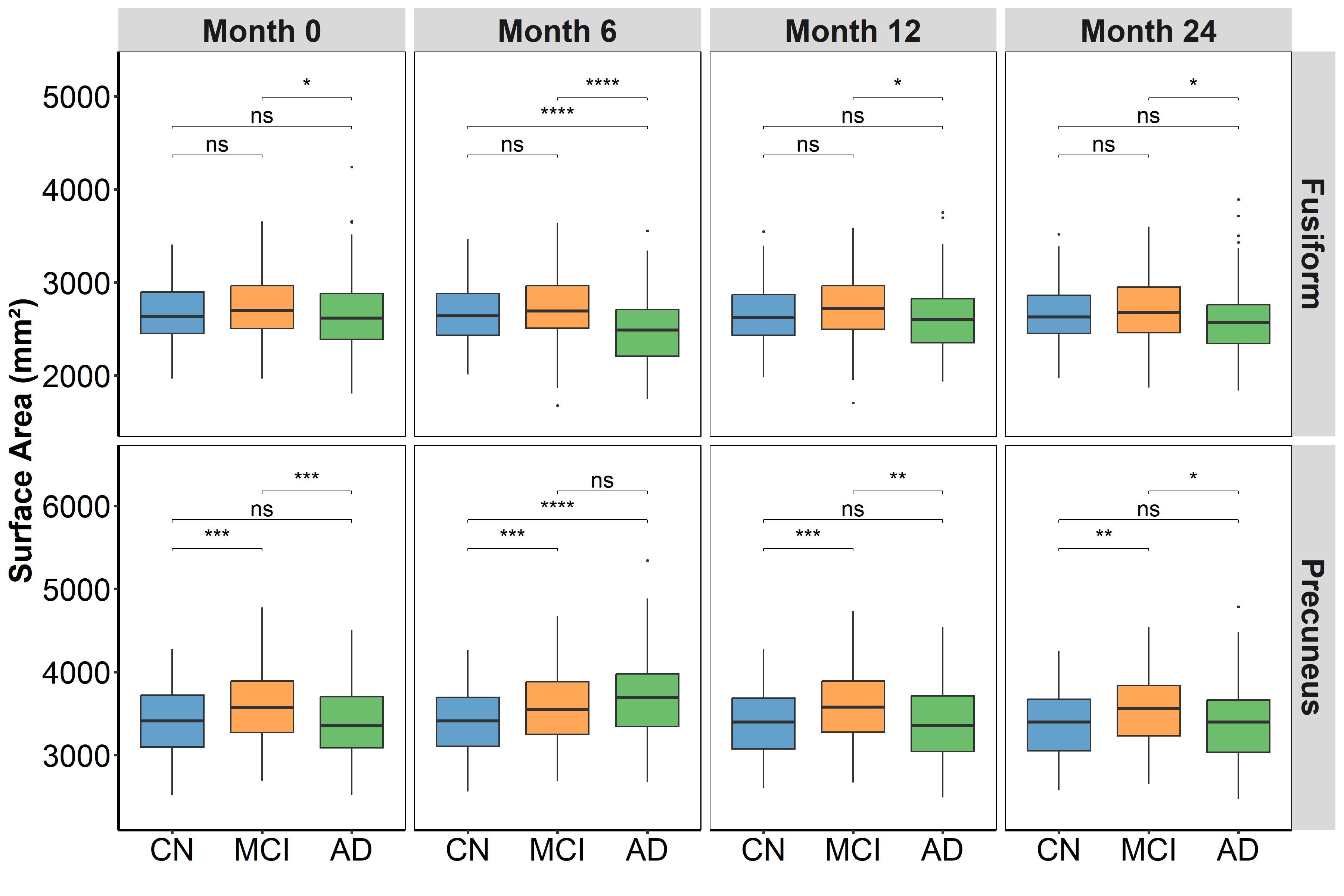

**Supp. Figure 15:** Surface area (SA) of representative samples from cognitive normal (CN), mild cognitive impairment (MCI), and Alzheimer's disease (AD) individuals at three separate time points (months 0, 6, and 12). A statistically significant difference (*: p ≤ 0.05; **: p ≤ 0.01; ***: p ≤ 0.001; ****: p ≤ 0.0001; ns: p > 0.05) is shown in a box plot based on the Mann-Whitney U test for SA of the left hemisphere of precuneus and fusiform gyrus.

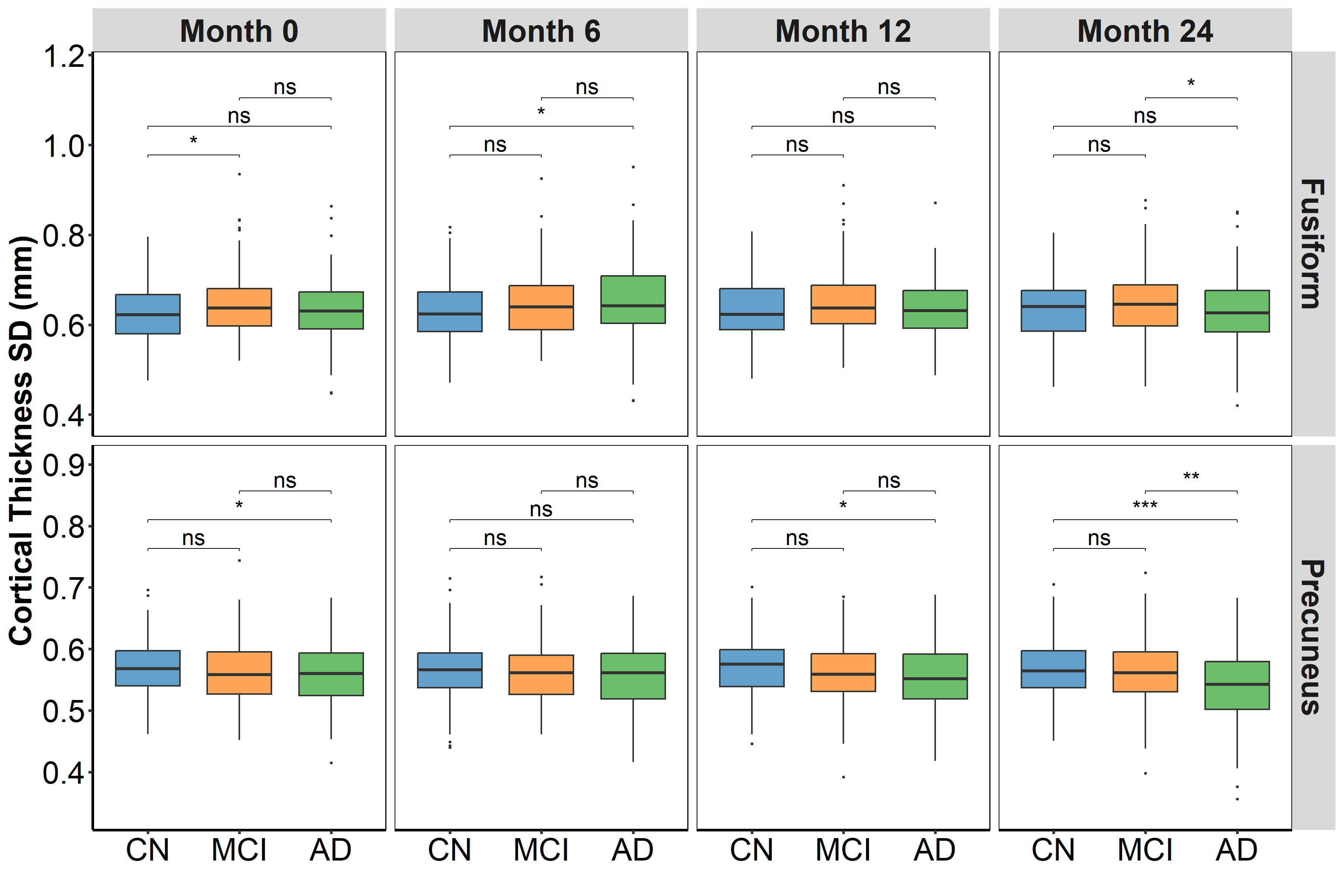

**Supp. Figure 16:** Cortical thickness Standard (CTS) of representative samples from cognitive normal (CN), mild cognitive impairment (MCI), and Alzheimer's disease (AD) individuals at three different time points (months 0, 6, and 12). A statistically significant difference (*: p ≤ 0.05; **: p ≤ 0.01; ***: p ≤ 0.001; ****: p ≤ 0.0001) is shown in a box plot based on the Mann-Whitney U test for CT of the left hemisphere of precuneus and fusiform gyrus.

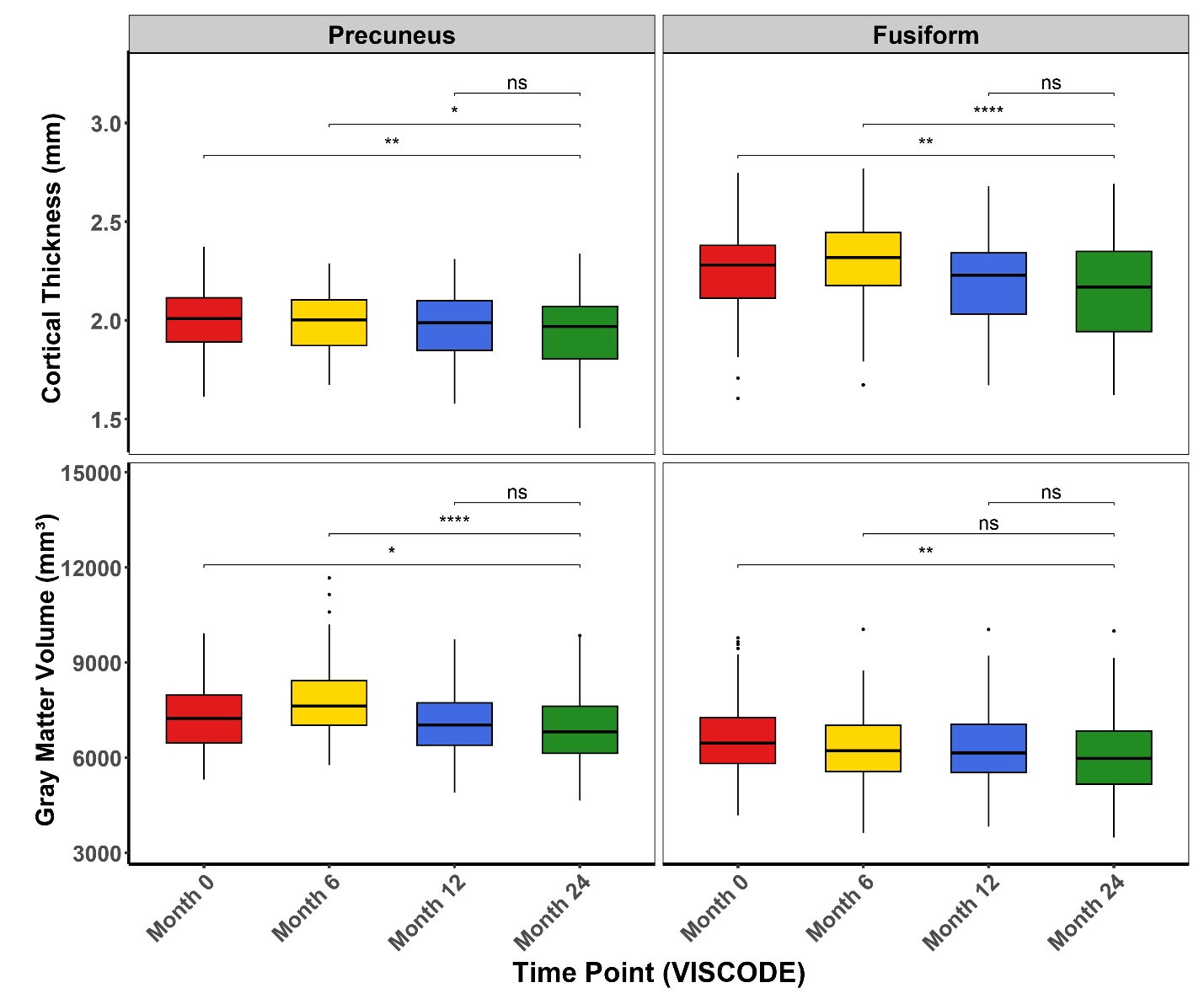

**Supp. Figure 17:** Boxplots illustrating GMV and CT of the fusiform gyrus and precuneus (left hemisphere) progression among AD individuals at 24-month time point (*: p < 0.05; **: p < 0.01; ***: p < 0.001; ****: p < 0.0001).

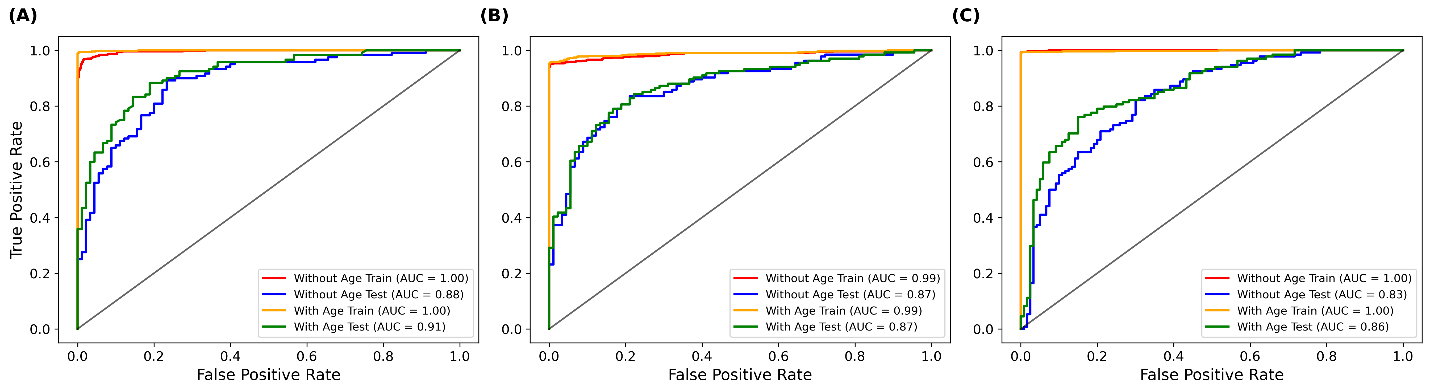

**Supp. Figure 18:** ROC plots for classification between (A) AD vs CN, (B) AD vs MCI, and (C) MCI vs CN using 18 radiomic features extracted exclusively from the fusiform gyrus (9 features from the left and 9 from the right hemisphere). Classifier models were developed both with and without the inclusion of age.

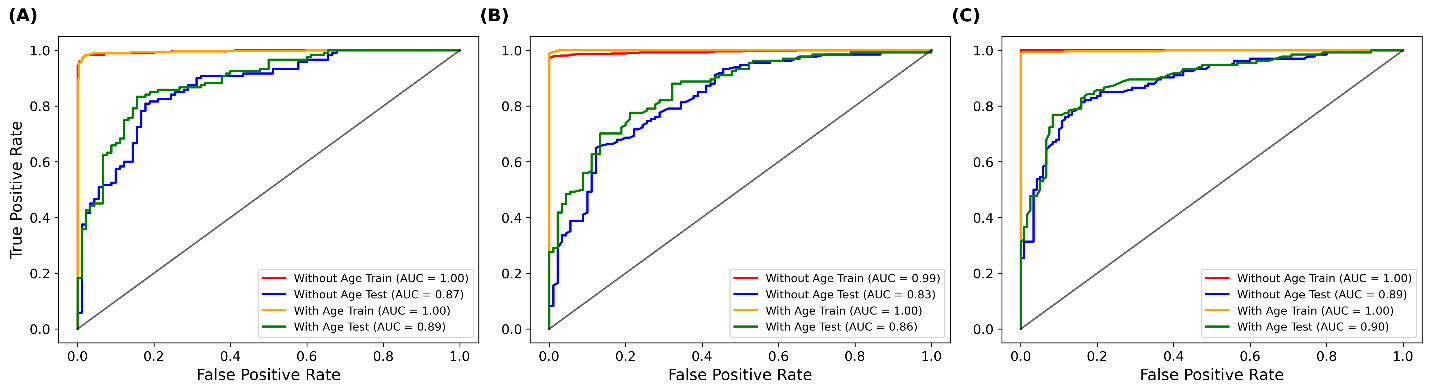

**Supp. Figure 19:** ROC plots for classification between (A) AD vs CN, (B) AD vs MCI, and (C) MCI vs CN using 18 radiomic features extracted exclusively from the precuneus (9 features from the left and 9 from the right hemisphere). Classifier models were developed both with and without the inclusion of age.
